# Early-life Exposome and Health-related Immune Signatures in Childhood

**DOI:** 10.1101/2025.03.21.25324385

**Authors:** Ines Amine, Augusto Anguita-Ruiz, Alicia Guillien, Xavier Basagaña, Mariona Bustamante, Eva Borràs, Marta Cirach, Audrius Dedele, Carlota Dobaño, Judith Garcia-Aymerich, Berit Granum, Regina Grazuleviciene, Juan Ramón González, Jordi Julvez, Hector Keun, Mónica López-Vicente, Rosemary McEachan, Gemma Moncunill, Mark Nieuwenhuijsen, Eduard Sabidó, Rémy Slama, Arthur Tenenhaus, Marina Vafeiadi, John Wright, Tiffany Yang, Wen Lun Yuan, Martine Vrijheid, Valérie Siroux, Léa Maitre

## Abstract

**Background:** Early-life environmental exposures are suspected to modify important immune processes related to child health. Yet, no study has investigated immunotoxicity in relation to the exposome and multiple health domains simultaneously.

**Methods:** Among 845 children (median age 8) from six European birth cohorts included in the Human Early-Life Exposome (HELIX) project, we identified immune signatures of a health score covering cardiometabolic, respiratory/allergic and neurodevelopmental health in children. Those signatures were identified from blood samples in three biological layers (white blood cell (WBC) composition, plasma proteins concentrations, DNA methylation of WBCs) using an advanced factorial analysis supervised on the child health score. Second, we estimated the association between the identified signatures and 91 pre- and postnatal environmental exposures.

**Results:** Three key immune signatures were associated with a better health score in children: a first protein signature characterizing a low inflammatory profile (R^2^=17%), a second protein signature characterizing a low inflammatory profile with balanced antiviral Th response (R^2^=2%), and a WBC signature characterizing an immuno-regulatory and naïve profile (R^2^=2%). In childhood, less exposure to indoor air pollutants, proximity to blue spaces and public transport, healthy dietary habits and higher social capital were associated with the three immune signatures related to a better health score (regression p-values<0.05). One signature was identified from DNA methylation, but was not significantly associated with the health score nor with the exposome.

**Conclusions:** These findings highlight the influence of early-life environmental exposures on key inflammatory processes associated with the cardiometabolic, respiratory and neurodevelopmental health of children.

**Highlights:**

– Three immune profiles linked to the child overall health were identified
– Those immune profiles were derived from multi-omics biomarkers in blood samples.
– 14 postnatal environmental exposures were associated with these immune profiles.
– It confirmed the environmental impact on key health-related inflammatory processes.

## Introduction

The exposome, encompassing all environmental exposures from conception onward, is a major determinant of human health across the lifespan (1,2). The early-life environment, including exposures to chemicals, atmospheric pollutants, natural spaces, and lifestyle factors, is known to influence the risk of chronic diseases and cancers later in life (3–6). While studies have traditionally focused on one or few specific environmental exposures, holistic approaches are now integrating the entire exposome (7). Those novel approaches were made possible with the development of large exposome datasets spanning hundreds of pre- and post-natal environmental exposures (8–15).

Alongside its clinical impacts, the exposome can alter fundamental biological processes, particularly those related to the immune system (16,17). These immune-related effects are of particular concern given the central role of the immune function in chronic diseases such as diabetes, obesity and respiratory disorders (18). Recently, the Lancet Commission on Planetary Health has recognized immunotoxicity as one of the most worrying and least studied consequences of environmental pollutants (19). Disruptions in immune function may contribute to shared pathways underlying a range of conditions across cardiometabolic, respiratory and mental health domains (20,21). Despite these insights, no exposome study has investigated immunotoxicity in relation to multiple health domains, with the aim of exploring the wide-ranging effects of environmental factors on health through immune profiles.

As immune processes span multiple biological layers (e.g. genetic, epigenetic, proteomics), the integration of multi-omics data could provide a more holistic characterization of the key mechanisms. While previous studies have explored the impact of the exposome on specific biological layers (22,23), few have used approaches integrating several biological layers (24,25). To overcome this limitation, the use of advanced statistical methods, capable of simultaneously analyzing multiple biological and clinical data layers, are of high interest. For example, methods that reduce dimensionality (e.g. latent components, clusters) are particularly promising for identifying complex, yet interpretable, immune processes (26–29).

This study, leveraging the Human Early Life Exposome (HELIX) cohort, seeks to address these gaps, by assessing the associations between a large range of early-life environmental exposures and multi-omics immune profiles linked to several child health domains.

## 1. Methods

The methodological workflow adopted for this project is illustrated in **Figure 1** and detailed below.

**Figure 1.**
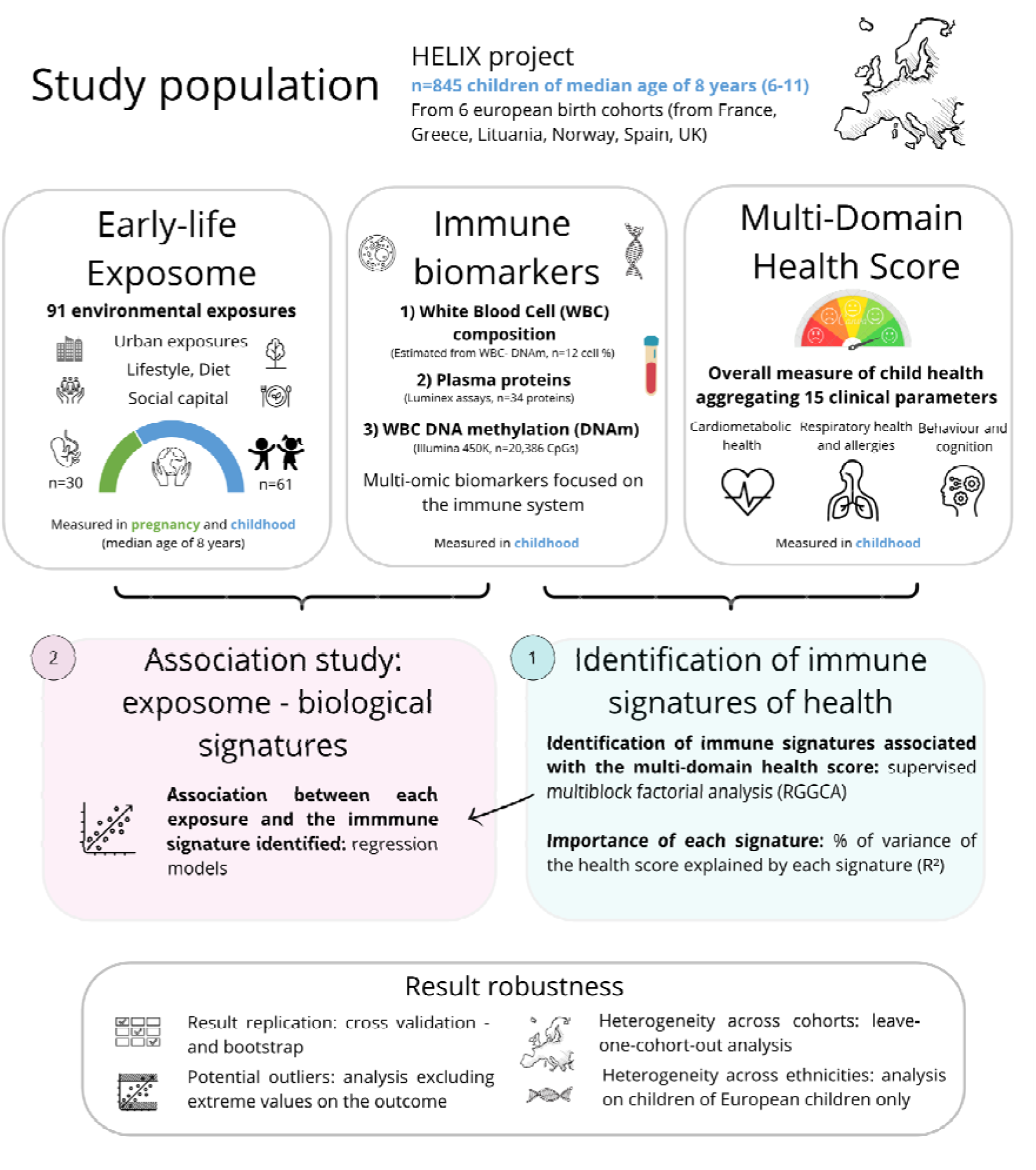
Project overview, This figure describes the methodology used for this project.

### a. Study population

This study is based on the HELIX project, including six population-based birth cohorts: Born in Bradford (BiB, UK, Wright et al. 2013), Étude des Déterminants pré et postnatals du développement et de la santé de l’Enfant (EDEN, France, Heude et al. 2016), Infancia y Medio Ambiente (INMA, Spain, Guxens et al. 2012), Kaunas Cohort (KANC, Lithuania, Grazuleviciene et al. 2011), The Norwegian Mother, Father and Child Cohort Study (MoBa, Norway, Magnus et al. 2016), and Mother-Child Cohort (RHEA, Greece, Chatzi et al. 2017). Initially, 32,000 mothers were recruited during pregnancy (2003-2009), from which 1,301 mother-child pairs were followed-up in 2014-2015 when the child was between 6 and 11 years, depending on the cohort (8,36). Standardized protocols were employed for biological sample collection, questionnaire administration, health examinations, and exposure characterization. The present study included 845 mother-child pairs with available data on the immune biomarkers and the outcome.

### b. Exposome

As detailed in **Table 1**, a total of 13 families of exposures were assessed in each mother-child pair, with 31 prenatal and 60 childhood exposures. They include outdoor exposures (outdoor air pollution, built environment, road traffic, surrounding natural spaces, meteorological data, water disinfections by products), indoor air pollutants, lifestyle (tobacco exposure, diet, physical activity, sleep, allergens) and socio-economic factors. Outdoor exposures were evaluated using geospatial methods and remote sensing at home and school addresses (37,38). Exposure to indoor air pollutants was assessed through predictive modeling based on a panel study of 150 mother-child pairs. Lifestyle factors, exposure to water disinfection by products and socio-economic factors were collected via questionnaires. More details on exposure assessment can be found in eMethods 1.

**Table 1.** Environmental exposures studied.

| Type of exposure | Exposures | Number of exposures Pregnancy | Number of exposures Childhood |
| --- | --- | --- | --- |
| **OUTDOOR & INDOOR EXPOSOME** |  |  |  |
| **Outdoor air pollution** | NO<sub>2</sub>, PM<sub>10</sub>, PM<sub>2.5</sub>, PM<sub>2.5</sub> absorbance | 4 | 4 |
| **Indoor air pollution** | NO<sub>2</sub>, PM<sub>2.5</sub>, PM absorbance, Benzene, TEX | 0 | 5 |
| **Meteorology** | Temperature, humidity, pressure, UV-vit D | 3 | 3 |
| **Surrounding natural spaces** | NDVI, presence of a major greenspace and bluespace | 3 | 3 |
| **Built environment** | Population density, building density, street connectivity, accessibility, facility richness, walkability, land use index | 9 | 9 |
| **Road traffic** | Traffic load on all roads and nearest road, traffic density on nearest road, inverse distance to nearest road | 4 | 4 |
| **Water DBPs** | THMs, brominated THMs, chloroform | 3 | 0 |
| Total of indoor and outdoor exposures for each period |  | 26 | 28 |
| **LIFESTYLE** |  |  |  |
| **Tobacco** | Active and passive smoking (pregnancy), child exposure to smoke (ETS) and parental smoking (childhood) | 1 | 2 |
| **Diet** | Food intakes (15 groups), food habits, supplements in folic acid, KIDMED score | 3 | 20 |
| **Sleep** | Average sleep | 0 | 1 |
| **Physical activity** | Moderate/vigorous activity, sedentary time | 0 | 2 |
| **Allergens** | Pet (all, dogs, cats) | 0 | 3 |
| Total of lifestyle exposures for each period |  | 4 | 28 |
| **SOCIO-ECONOMIC** |  |  |  |
| **Economic status** | Family Affluence Score (FAS) | 0 | 1 |
| **Social capital** | Family contact, participation in organizations, family size, maternal stress | 1 | 3 |
| Total of socio-economic exposures for each period |  | 1 | 4 |

Abbreviations: KIDMED: Mediterranean Diet Quality Index for children, NO<sub>2</sub>: Nitrogen dioxide, PM:
Particulate matter, TEX: Toluene, Ethylbenzene, and Xylene.

### c. Immune biomarkers

Three biological layers were analyzed from blood samples in childhood (6-11 years): **1) White Blood Cell (WBC)** composition estimated from DNA methylation measured in buffy coat, **2) plasma proteins (Prot)** concentrations and **3) DNA methylation (DNAm)** of white blood cells (genome-wide) in buffy coat. **Table 2** details the biomarkers studied and the measurement methods used, with more details in eMethods 2. Briefly, plasma proteins were estimated by Luminex using the Cytokines 30-plex, the Apolipoprotein 5-plex, and the Adipokine 15-plex. DNAm of WBC was assessed in buffy coat using Illumina 450K (genome-wide), from which WBC composition was derived using the Houseman method (39). For the study of DNAm, pre-processing included addressing technical issues such as batch effects and WBC composition through residualization analysis (40,41), filtering out CpGs with high technical variability (42) and those with no association with the exposome based on the study of Maitre (25). A total of 20,386 CpG sites were selected from the 480,071 sites initially measured.

**Table 2.**
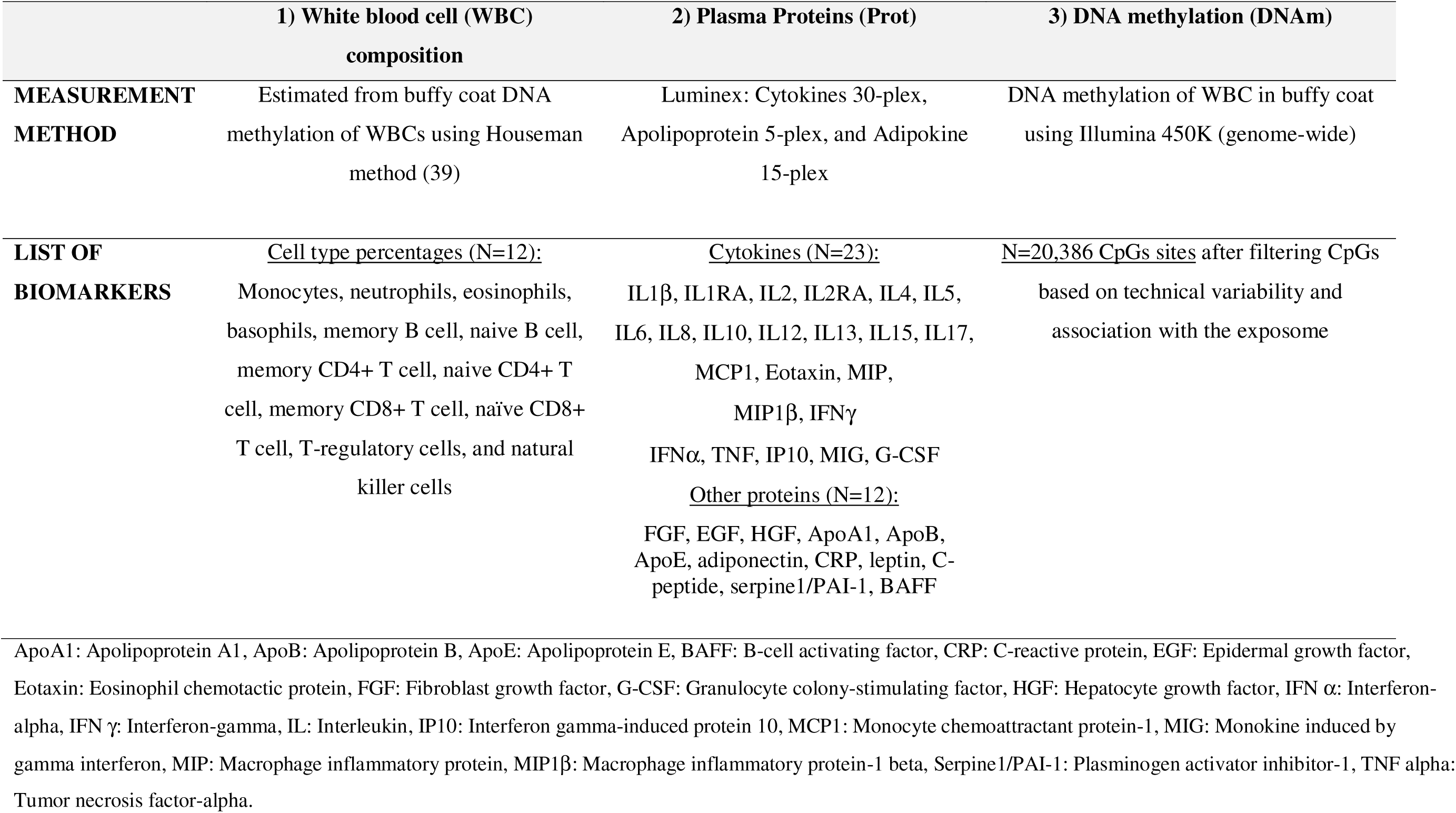
Immune biomarkers studied.

### d. Multi-Domain Health score

We integrated pre-clinical and clinical data through a multi-domain health score, thereafter called “health score”, developed in a previous HELIX study (Amine et al. 2023). It is derived from three subscores (respiratory, cardiometabolic, and neurodevelopmental) aggregating overall fifteen health parameters (more details in eMethod 3). By design, a higher health score indicates better health status in children. This score is low-to-moderate for children who exhibit low-to-moderate health in cardiometabolic, respiratory/allergy and neurodevelopmental domains simultaneously, as well as for children who are severely affected in one health domain while being unaffected or poorly affected in the other two.

### e. Covariates

Confounders adjusted for in the analysis included cohort, child age (continuous), sex, ethnicity (European vs non-European origin), maternal education (low, medium, high), breastfeeding duration (<11 weeks, 11-35 weeks, >35 weeks), pre-pregnancy BMI (kg/m2), maternal age (years), season of birth (winter, spring, summer, autumn), time since last meal (analyses on plasma proteins only), and hour of blood collection (analyses on plasma proteins only). Exposures, immune biomarkers and the health scores were pre-corrected on these covariates using a residualization approach (40,41).

### f. Statistical analysis

Exposures and covariates were transformed to reach normality when needed (see eTable 1), and imputed using chained equations (44) (first imputed dataset used), as detailed in eMethod 4. Continuous exposures were centered and standardized by the interquartile range (see eTable 1).

In the first step, immune signatures predictive of the multi-domain health score were identified using a multi-block model (see more details in eMethods 5). Regularized Generalized Canonical Correlation Analysis (RGCCA) (*RGCCA* R package), an advanced multi-block supervised factorial analysis model, was applied to estimate such signatures, also called “latent components”, in each data block (WBC, proteins and DNAm) (45). The optimal number of signatures, and a sparsity parameter used for DNAm, were estimated based on cross-validation. Evaluation of the model was performed using cross validation: we reported the R^2^ and p-value based on linear regression models to assess the ability of the immune signatures (individually or together) to predict the health score and its sub-scores (cardiometabolic, respiratory, and neurodevelopmental). Interpretation of the signatures was based on the contribution of each immune biomarker given by the loadings, i.e. the correlation between a biomarker and the signature, and 95% intervals were estimated by bootstrap.

In a second step, associations between environmental exposures and the identified immune signatures were assessed using univariate linear regression models.

To address sex-specific associations, the analysis was further stratified on sex. In addition, robustness checks were performed through sensitivity analyses, including leave-one-cohort-out, removal of the 4% extreme values on the outcome, analysis restricted to children with European-origin, and analysis using DNAm levels uncorrected for WBC composition. We also compared the results to a more traditional “Meet in the middle” approach (46), which aims to identify the immune biomarkers simultaneously associated with environmental exposures and the child health scores, based on regression models (see eMethod 5). Analyses were done with R version 4.2.1, and codes are publicly available on GitLab (47).

## 2. Results

### a. Description of the population

The study population comprised 47% girls, and children had a median age (Q25; Q75) of 8 years (6.5; 8.9) at follow-up (eTable 2). The median maternal age at childbirth was 31 years, with 51% of mothers having a high degree of education. Description of the immune biomarkers (eFigure 1) and the multi-domain health score is available in supplementary materials (eFigure 2, eTable 2 and 3), and exposure levels were previously described (37,43).

**Figure 2.**
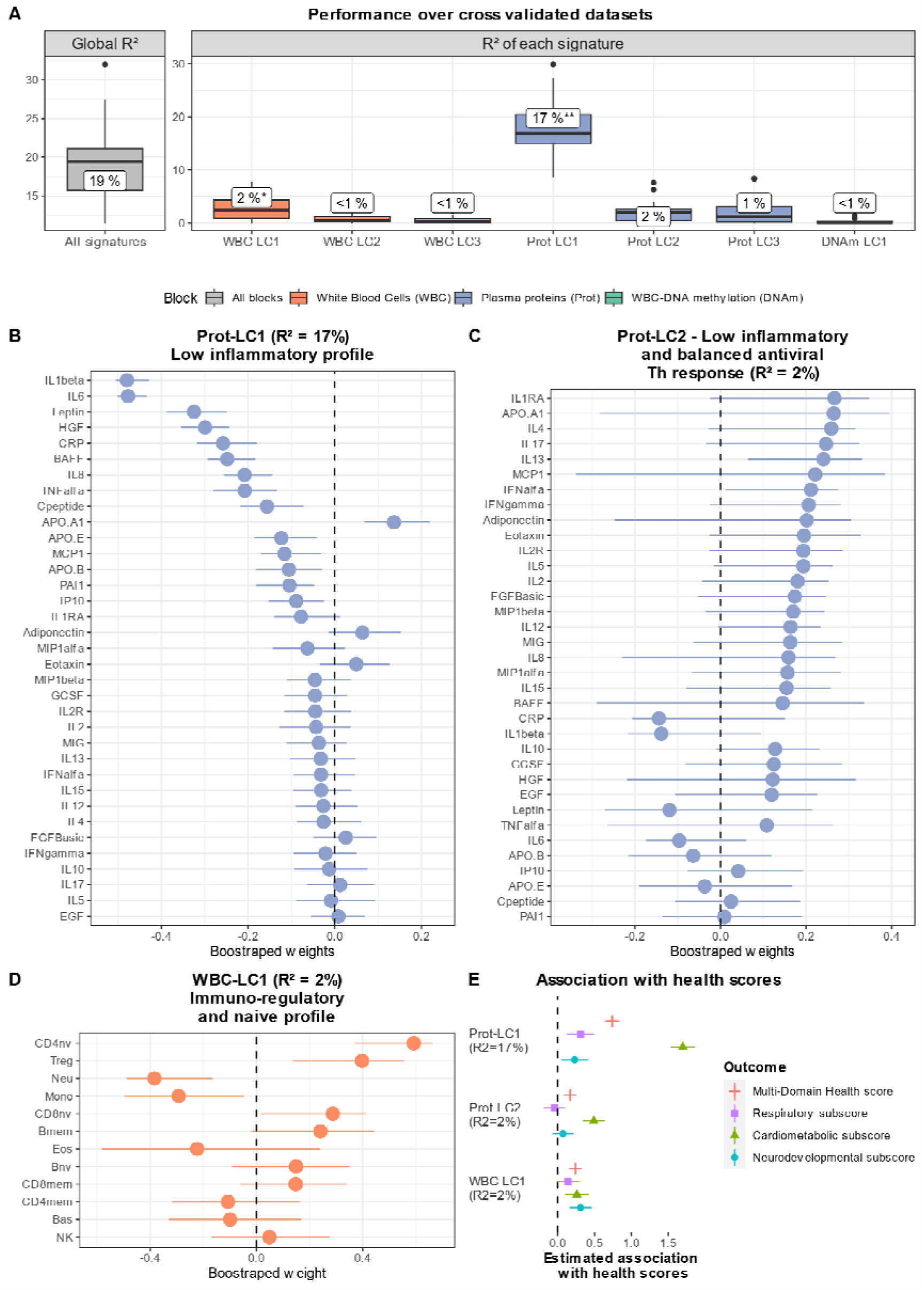
Immune signatures estimated using a multi-block model, This figure describes the immune signature (latent components) estimated using a multi-block model (Regularized Generalized Canonical Correlation Analysis). It includes the distribution of the performance indicators across the cross validated datasets (5-fold repeated 5 times) (panel A), as well as biomarker loadings for the main immune signatures which are Prot-LC1 (panel B), Prot-LC2 (panel C) and WBC-LC1 (panel D) and their association with the health scores (panel E). For panel A, Rsquared (R2) and p-values have been obtained by fitting a linear regression between the health score and the given signature (or all signature for “Global R2”), with * or ** meaning that the p-value of association was below 0.05 or 0.01, respectively. For panels B to D, the point represents the biomarkers loading, i.e. the correlation between a given biomarker and a given signature, and 95% confidence intervals were computed using bootstrapping. Panel E represent the estimated coefficient of association and 95% confidence interval between Prot-LC1 and WBC-LC1 with the different health scores, obtained by linear regressions. Abbreviations: ApoA1: Apolipoprotein A1, ApoB: Apolipoprotein B, ApoE: Apolipoprotein E, BAFF: B-cell activating factor, Bas: Basophils, Bmem: B memory, Bnv: B naïve, CD4mem: CD4 memory, CD4nv: CD4 naïve, CD8mem: CD8 memory, CD8nv: CD8 naïve, CRP: C-reactive protein, EGF: Epidermal growth factor, Eos: Eosinophils, Eotaxin: Eosinophil chemotactic protein, FGF: Fibroblast growth factor, G-CSF: Granulocyte colony-stimulating factor, HGF: Hepatocyte growth factor, IFN α: Interferon-alpha, IFN gamma: Interferon-gamma, IL: Interleukin, IP10: Interferon gamma-induced protein 10, MCP1: Monocyte chemoattractant protein-1, MIG: Monokine induced by gamma interferon, MIP: Macrophage inflammatory protein, MIP1beta: Macrophage inflammatory protein-1 beta, Mono: Monocytes, Neu: Neutrophils, NK: Natural Killers, Prot: Proteins, Serpine1/PAI-1: Plasminogen activator inhibitor-1, TNF α: Tumor necrosis factor-alpha. Treg: T regulatory, WBC: white blood cell.

### b. Identification of immune signatures of the health score using a multi-block model

Based on cross validation, the multi-block model identified seven Latent Components (LC), representing seven health-related immune signatures: one for DNA methylation (DNAm), three for plasma proteins (Prot) and three for WBC composition, as shown in **Figure 2**. The signatures were poorly correlated with each other (maximum rho = 0.2, see eTable 4). Together, these immune signatures explained 19% of the multi-domain health score (cross validated R^2^). By design, all signatures were associated with a better multi-domain health score.

The first protein signature (Prot-LC1) best predicted the multi-domain health score (cross-validated R^2^=17%), followed equally by the second protein signature (Prot-LC2) and the first WBC signature (WBC-LC1) (R^2^=2% each) (see Figure 2). The other signatures (Prot-LC3, WBC-LC2, WBC-LC3, DNAm-LC1) were excluded because of their weak associations with the child health score (cross-validated p-value > 0.1), but their description can be found in the supplementary materials (eFigure 3).

**Figure 3.**
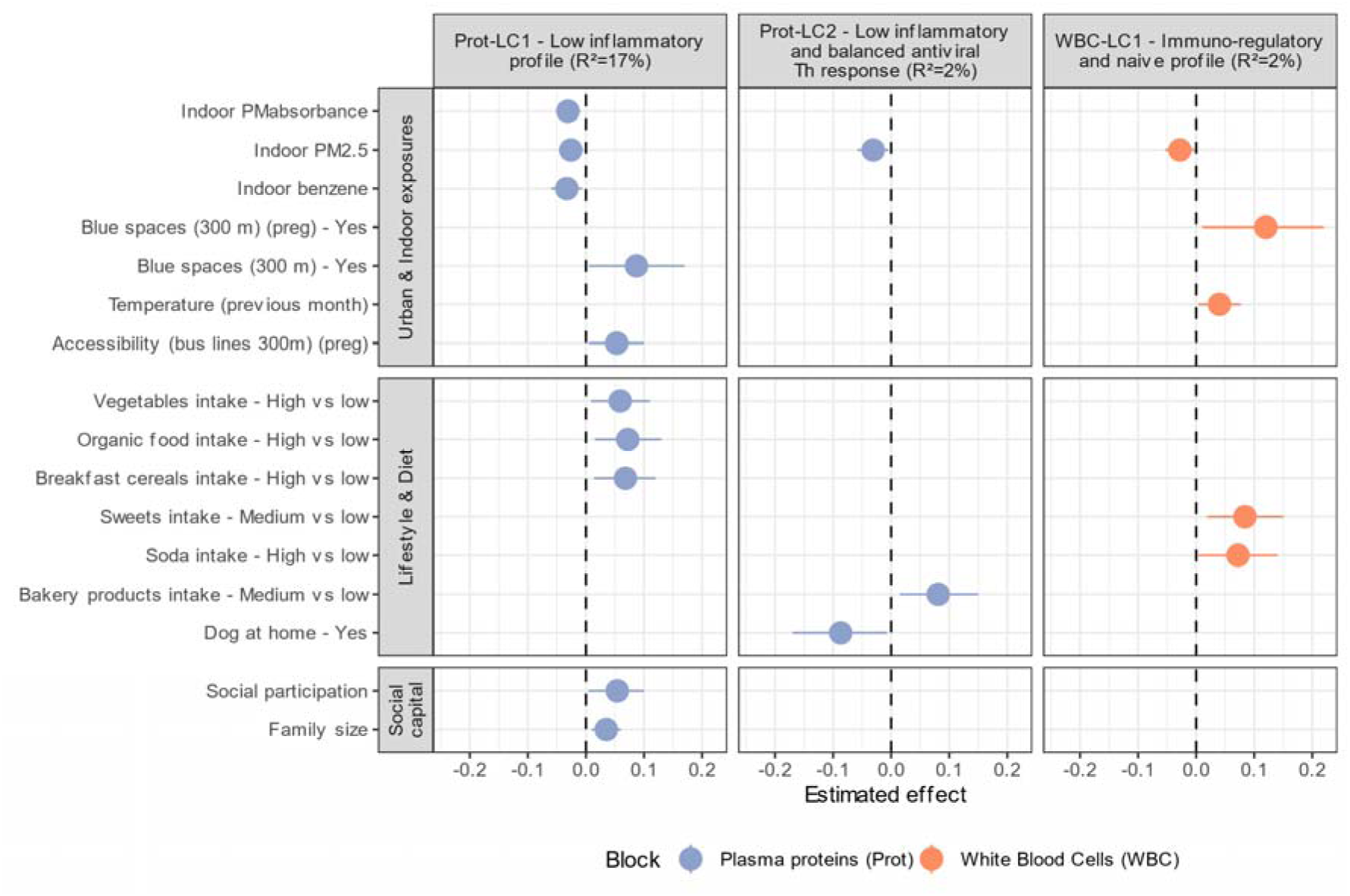
Significant associations between exposures and the main immune signatures, Significant associations between exposures and the main immune signatures estimated using the multi-block model (RGCCA) in the study population (n=845), with a p-value threshold of 5%. Exposures labeled with “(preg)” refer to pregnancy-related exposures, while the others were measured during childhood (ages 6-11 years). The R^2^ values represent the cross-validated R^2^ from fitting a linear regression between the respective immune signature and the health score. Abbreviation: PM: particulate matter, Prot-LC1: first protein latent component, Prot-LC2: second protein latent component, WBC-LC1: first white blood cell latent component.

The main contributors to Prot-LC1 were IL1β, IL6, leptin, Hepatocyte Growth Factor (HGF) and C-Reactive Protein (CRP), all with negative loadings. Therefore Prot-LC1 was labeled as a “Low inflammatory profile” (see Figure 2 and and supplementary table *Online_Immune_Signatures*). The signature Prot-LC2 was characterized by high levels of IL1RA, IL4, IL17, IL13 and IFNα, thus it was labeled as a “Low inflammatory and balanced antiviral Th response”. Lastly, the signature WBC-LC1 was characterized by high proportions of T regulatory cells, CD4 naïve, CD8 naïve, and lower proportions of neutrophils, monocytes and eosinophils, therefore it was labeled as an “Immuno-regulatory and naïve profile”. As shown in Figure 2 and detailed in eTable 5, Prot-LC1 and WBC-LC1 were significantly associated with all subscores (cardiometabolic, respiratory, neurodevelopmental), and Prot-LC1 and Prot-LC2 were more strongly associated with the cardiometabolic subscore than with the neurodevelopmental and respiratory subscores.

### c. Association between the exposome and the health-related immune signatures

As shown in **Figure 3**, several environmental factors were associated with immune signatures related to a better health score (see also eTable 6). Among outdoor and indoor exposures, childhood indoor exposure to particulate matters with a diameter less than or equal to 2.5 micrometers (PM_2.5_), PM_2.5_ absorbance and benzene was associated with a decrease in Prot-LC1, with indoor PM_2.5_ also associated with decreased WBC-LC1 and Prot-LC2. On the other hand, surrounding blue spaces (size greater than 5,000 m² in a 300m buffer near residence) during pregnancy and childhood were associated with higher WBC-LC1 and Prot-LC1, respectively. Higher ambient temperature (the month preceding the visit) and better access to transports (bus lines in a 300m buffer) during pregnancy were associated with higher WBC-LC1 and Prot-LC1, respectively.

Regarding lifestyle and diet, high intakes of vegetable, organic food, and breakfast cereals (vs low intake) in childhood was associated with higher Prot-LC1. Moderate intake of sweets (*vs.* low intake) and high intake of soda (*vs.* low intake) were associated with higher WBC-LC1. Moderate intake of bakery products was associated with higher Prot-LC2. The household presence of a dog in childhood was associated with lower Prot-LC2.

Finally, two social factors were associated with higher Prot-LC1, namely the child social participation in an organization (vs none) and the number of people living in the house.

### d. Supplementary analyses

The multi-block model revealed similar signatures for boys and girls, except for Prot-LC2, whose loadings displayed significant instability in females (see eFigure 4). In addition, results from the sensitivity analyses reinforced those from the main analysis (see eFigure 5-6 and supplementary table *Online_Immune_Signatures*), with similar signatures identified. Using DNAm values uncorrected for WBC increased the R^2^ of DNAm-LC1 to 2.7% (*vs*. 0.1% in the main analysis), with a built signature strongly correlated with WBC-LC1 (rho=0.9) (see eTable 7 and 8). Finally, the “Meet-in-the-Middle” analysis, based on univariate models, highlighted a similar set of biomarkers to the multi-block model (see the supplementary file *Online_Meet_in_the_Middle*).

## 3. Discussion

This study uniquely explores how environmental exposures influence key health-related immune signatures in children, identified using an advanced multi-block model (RGCCA). It simultaneously integrated exposome data, multi-omic biomarkers focused on the immune system, and a multi-domain health score covering the child cardiometabolic, respiratory and neurodevelopmental health. We identified three immune signatures associated with a better health score in children: Prot-LC1 (low inflammatory profile), WBC-LC1 (immuno-regulatory and naïve profile), and Prot-LC2 (low inflammation and balanced antiviral Th response). Among childhood exposures, better indoor air quality, proximity to blue spaces, healthy dietary intakes, and a strong social capital were associated with immune signatures related to a better health score. Although a DNA methylation signature was also identified, it showed no significant association with the health score or the exposome. These findings highlight the influence of several environmental factors on key inflammatory processes, which hold significant clinical relevance for cardiometabolic, respiratory and neurodevelopmental health in children.

### a. Main findings

Using RGCCA, an advanced dimension reduction technique, we identified three key immune signatures—Prot-LC1, Prot-LC2, and WBC-LC1—associated with better outcomes across three key health domains in children (cardiometabolic, respiratory, neurodevelopment). Prot-LC1, the primary signature (R^2^=17%), reflects a low inflammatory profile, characterized by reduced levels of pro-inflammatory cytokines (IL1β, IL6, TNF), CRP, and HGF, the latter suggesting a tissue repair response to inflammation-induced stress (48,49). The inclusion of leptin highlights the interplay between metabolism and inflammation, with metabolic disorders (e.g., obesity) driving cytokine production by fat cells (e.g., IL1β, IL6) (50,51). The secondary signatures, WBC-LC1 and Prot-LC2 (R^2^=2% each), further emphasize the key role of inflammatory regulation, as evidenced by the contribution of regulatory T cell and myeloid cells in WBC-LC1 (52,53) and anti-inflammatory cytokines (IL1RA, ILR2) in Prot-LC2. In addition, these signatures reflect immune responses to viral infections, with elevated IFNα in Prot-LC2 and high naive T cells in WBC-LC1, suggesting the presence and absence of each sub-score (cardiometabolic, respiratory/allergic, and neurodevelopmental) highlighting the central role of inflammation across various health domains (54,55).

The results of this study suggest that better indoor air quality, the absence of a dog, and higher ambient temperature, may significantly improve the health-related immune signatures identified. Better indoor air quality (PM_2.5_, PM_2.5_ absorbance, benzene) was associated with favorable immune signatures at both protein and WBC levels, likely due to the PM effects on oxidative stress (56) and the link between oxidate stress and inflammation (57). Moreover, the absence of a dog was associated with better Prot-LC2, suggesting balanced Th response in absence of allergens, although the literature on this matter remains unclear (58). Lastly, higher ambient temperature in the month preceding the health visit was associated with better WBC-LC1, possibly reflecting decreased respiratory infections during warmer seasons (59,60), as evidenced in WBC-LC1 by higher proportions of naive versus memory cells.

In addition, multiple types of child food intakes were associated with the immune signatures. A healthy diet, rich in vegetables, organic foods and including breakfast, was associated with higher Prot-LC1 (i.e. better health), reflecting their benefits for inflammation regulation and metabolic health (61,62). On the other hand, moderate sweet and bakery product intakes and high soda consumption were associated with higher WBC-LC1 or Prot-LC2. While moderate sugar intake may have some benefits, the association with high soda intake, including artificially sweetened options, warrants further investigation (63,64).

Lastly, stronger social capital and proximity to natural spaces and public transports, were associated with immune signatures (Prot-LC1 and WBC-LC1) related to better health. Protective associations were found with the family size and the child participation in organizations, possibly reflecting the importance of a good social network in childhood for stress reduction and inflammation regulation, and consequently to a good overall health (65,66). Similarly, surrounding blue spaces and public transport near the residency appeared to be associated with healthier immune signatures, likely by promoting social interactions and encouraging physical activity (67,68).

While the exposome is suspected to influence epigenetic processes (69,70), the signature from DNA methylation of WBCs showed no significant association with the health score (cross-validated p > 0.1) or environmental exposures. Several hypotheses may explain this: 1) the environmental exposures used to pre-select CpGs may not affect clear epigenetic mechanisms due to low intra-block correlation, 3) the sample size was too small for replication using cross-validation, and 4) correction of DNA methylation for WBC effects may have removed important variability. Regarding the later hypothesis, a sensitivity analysis showed improved R^2^ when WBC effects were retained, with new signatures from DNAm and WBC highly correlated. These results confirm the key role of cell composition on health, which might be more direct than epigenetic mechanisms alone.

### b. Strength and weaknesses

This study has several strengths, including the analysis of numerous exposures, immune biomarkers, and health outcomes, providing a comprehensive view of immunotoxicity in relation to the exposome and multiple health domains. By integrating multi-omic data with the RGCCA method, an advanced multi-block dimension reduction technique, we were able to identify complex yet interpretable immune processes. We have made the codes used in this project publicly available, providing a valuable resource for future exposome-omics studies (Amine 2025). Moreover, we ensured the strong clinical relevance of these immune processes by supervising the model on a novel health score that integrates clinical and pre-clinical parameters of cardiometabolic, respiratory and neurodevelopmental health in children.

However, we recognize that our study has some limitations. The multi-domain health score aims to study simultaneously three health areas; thus, it cannot identify immune signatures specific to an individual health domain. Additionally, WBC analysis relied on estimated cell proportions by using a validated model, rather than measured counts (39). Similarly, exposure to indoor air pollutant was assessed using prediction models from a subset of participants (n=150), warranting cautious interpretation. Lastly, the cross-sectional study design limits causal inferences, with exposures (at the exception of those from pregnancy), immune biomarkers, and outcomes measured simultaneously in childhood. To avoid potential reverse causality due to this design, we preferred not to analyze chemical pollutants, which were measured in HELIX in the same biological samples as the immune biomarkers.

## Conclusion

This novel study investigated associations between the early-life exposome and health-related immune signatures in children. The findings suggest that better indoor air quality, proximity to natural spaces, healthy dietary habits, and strong social capital are associated with reduced and better-regulated inflammation, at the protein and white blood cell levels. Overall, these results underline the importance of these environmental factors in mitigating immunotoxicity related to multiple areas of health in children.

## Supporting information

Online table 1 - Immune signatures

Online table 2 - Meet in the middle

## Data Availability

The access guidelines for HELIX data is available with the data inventory in this website: http://www.projecthelix.eu/data-inventory. The document describes who can apply to the data and how, the timings for approval and the conditions to data access and publication.

## Acknowledgment

We are grateful to all the participating families, practitioners, schools and researchers in France, Greece, Lithuania, Spain, Norway and UK who made this study happen. This project is only possible because of their enthusiasm and commitment.

