## Supplementary materials for "Early-life Exposome and Health-related Immune Signatures in Childhood"

### Supplementary figures

#### eFigure 1 - Distribution of the immune biomarkers (plasma proteins and white blood cell composition)

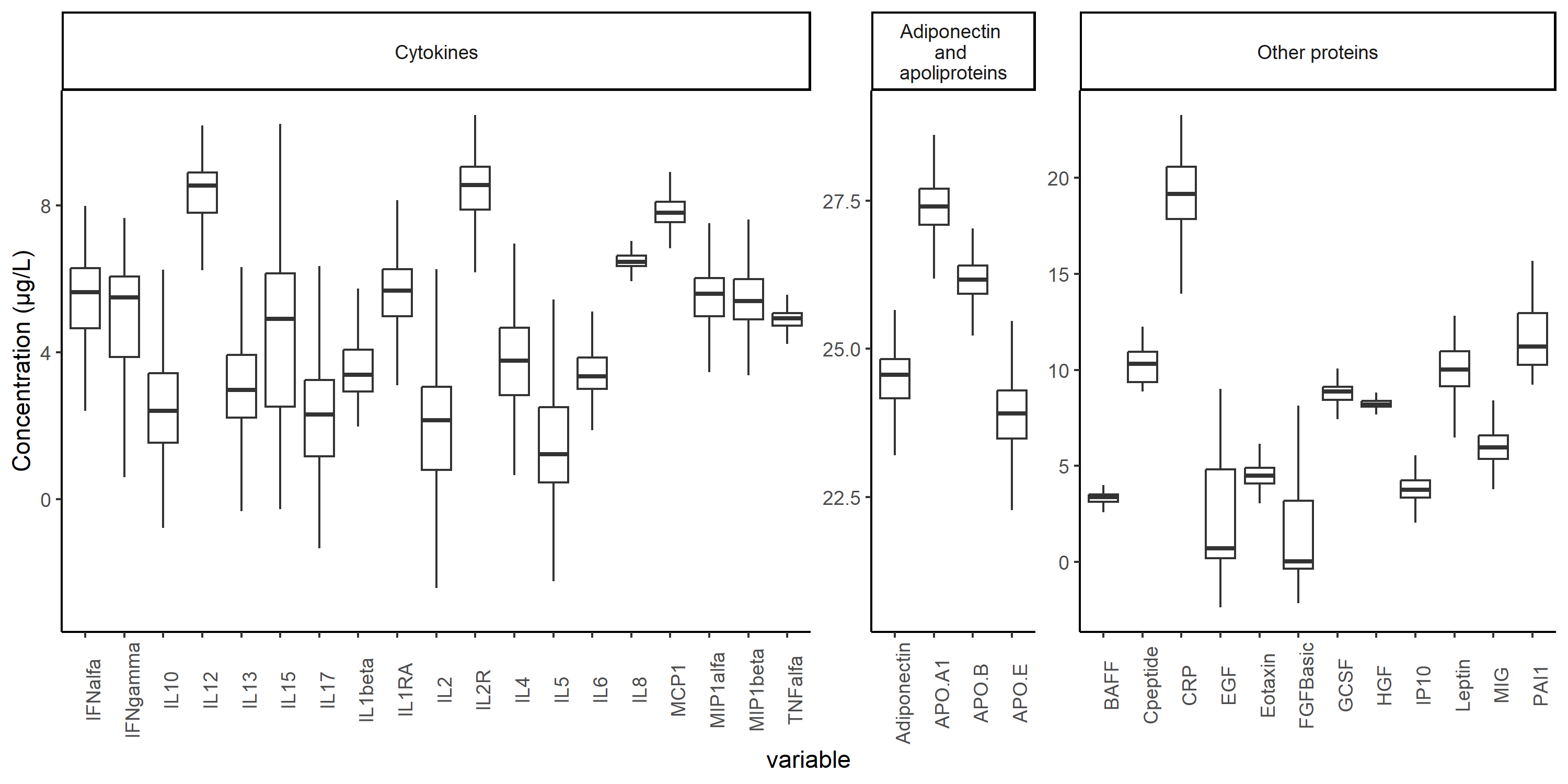

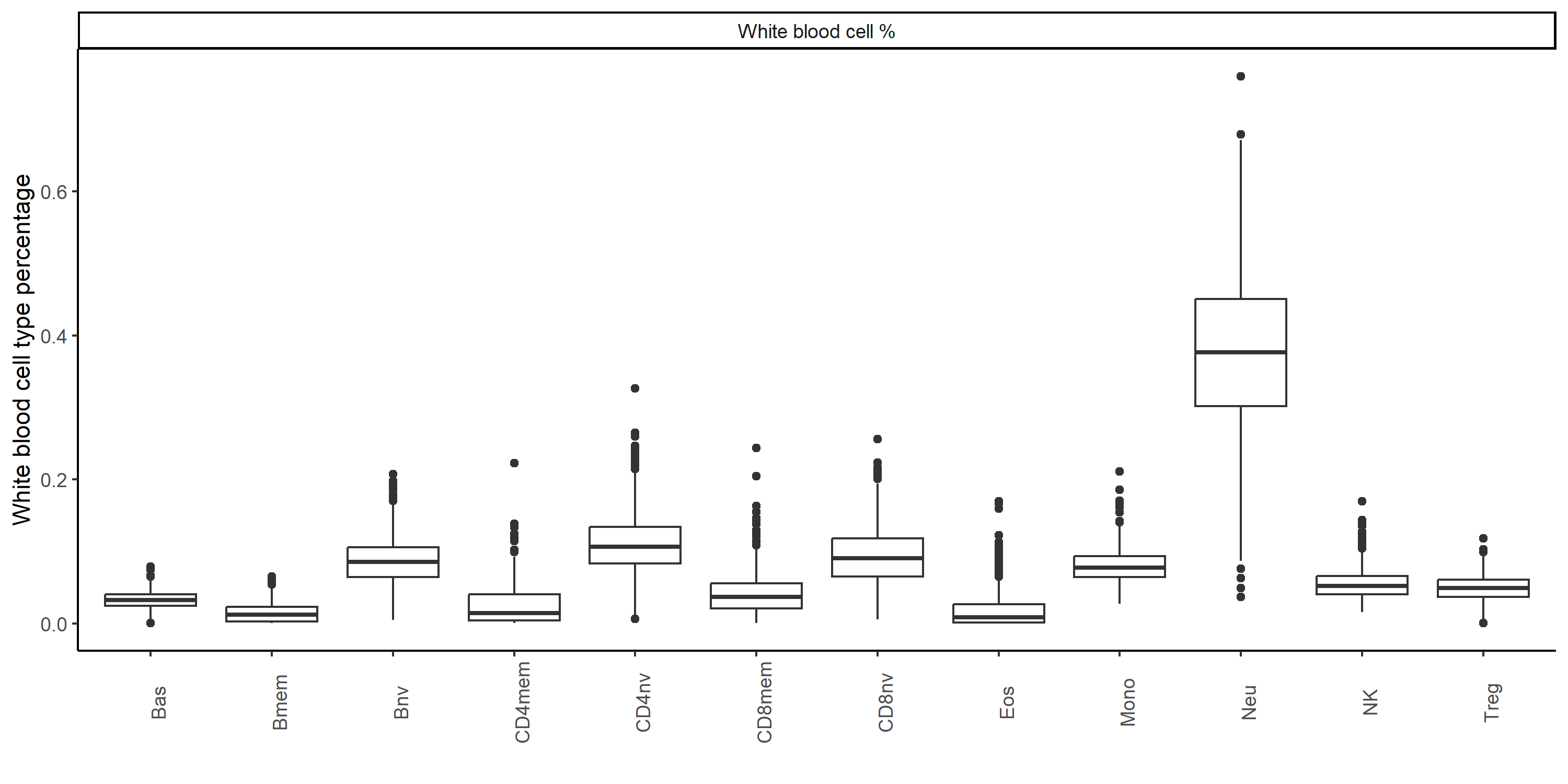

The boxplots show the distribution of proteins and white blood cell percentages in the study population, after pre-treatment (n=845).

#### eFigure 2 - Distribution of the health score in the whole population and by cohort

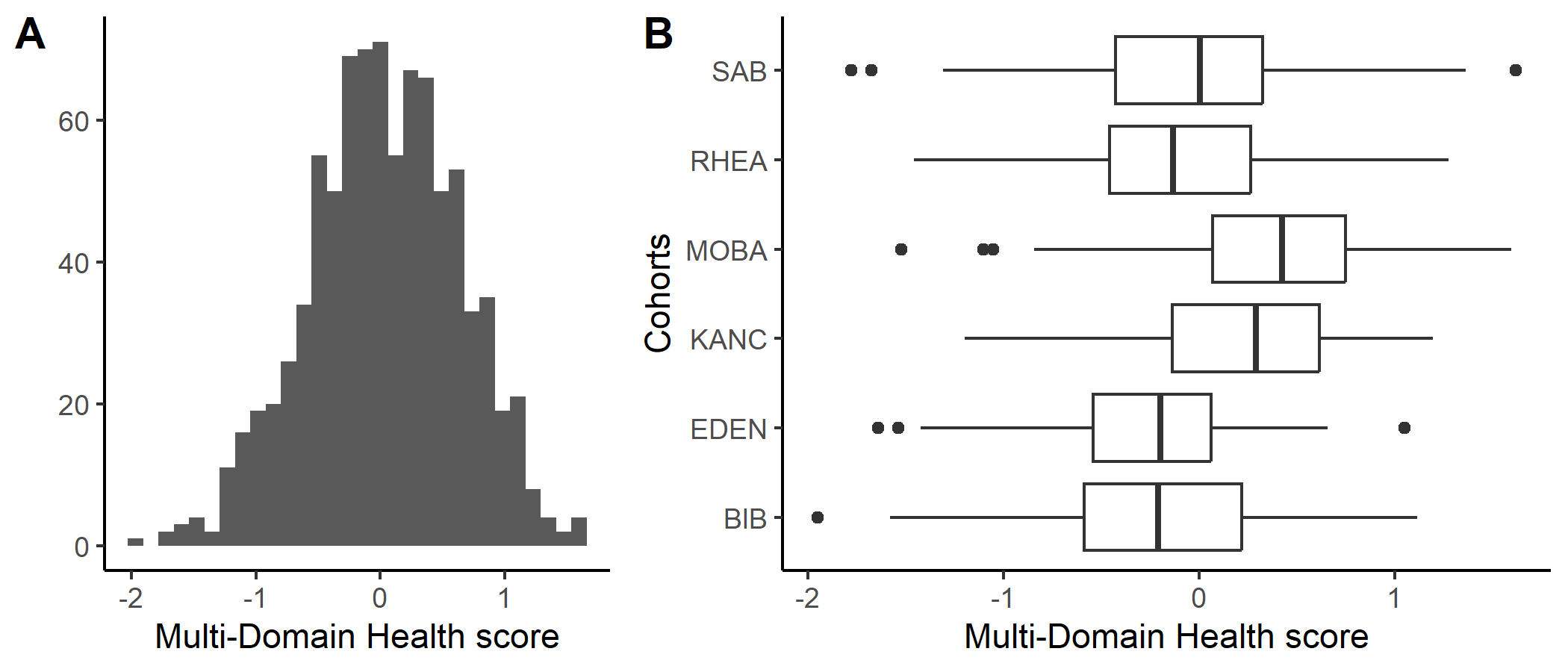

This figure shows the distribution of the multi domain health score in the whole population (panel A) and each cohort site (panel B).

#### eFigure 3 – Description of the immune signatures WBC-LC2, WBC-LC3, Prot-LC3 and DNAm-LC1, not kept for later analyses

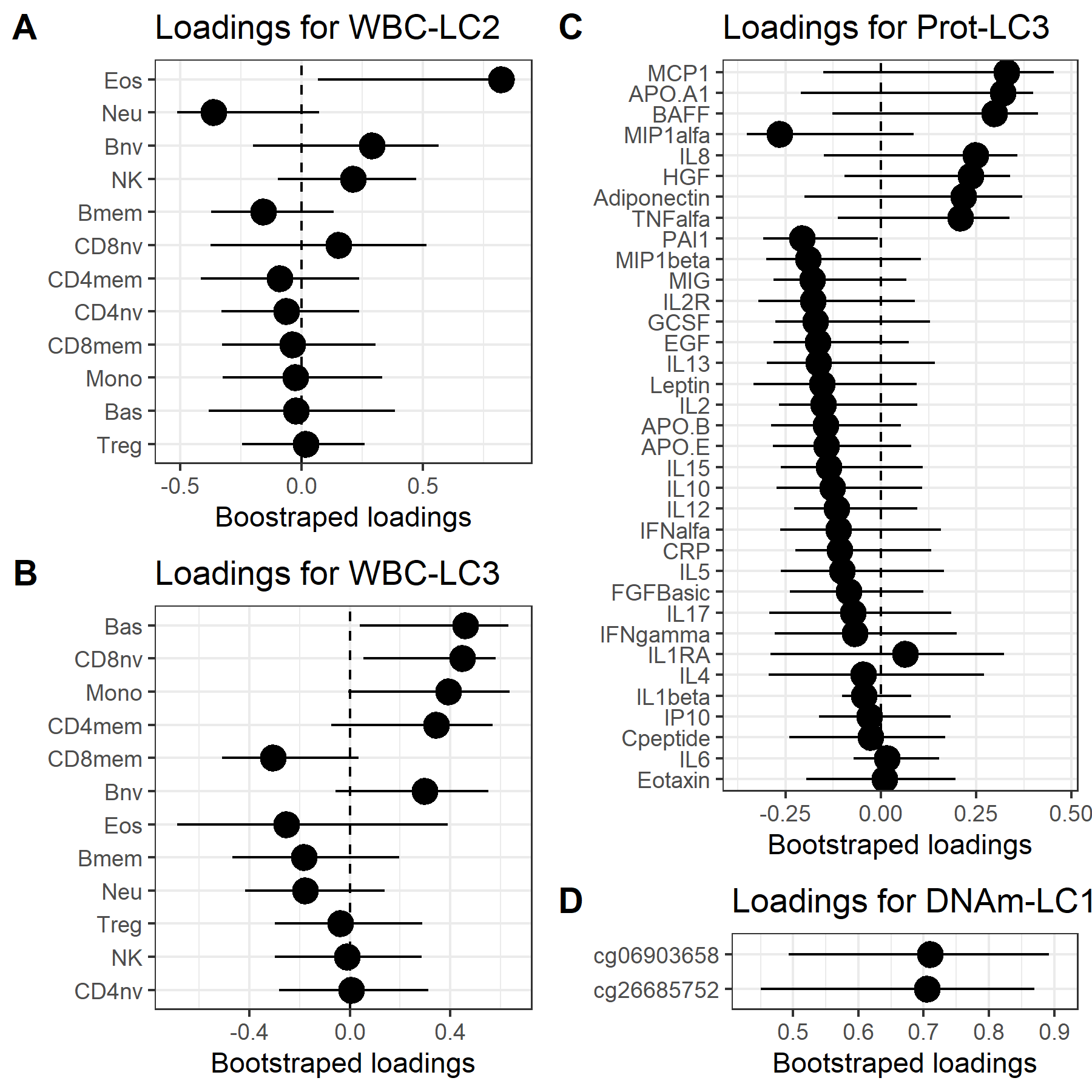

This figure describes biomarker loadings for components not described in the main paper because of low R2 or unstability. WBC-LC2 (panel A), WBC-LC3 (panel B), Prot-LC2 (panel C) and DNAm-LC1 (panel D). The point represents biomarkers loadings, i.e. the correlation between each biomarker and a given component, and 95% confidence intervals were computed using bootstraping.

ApoA1: Apolipoprotein A1, ApoB: Apolipoprotein B, ApoE: Apolipoprotein E, BAFF: B-cell activating factor, Bas: Basophils, Bmem: B memory, Bnv: B naïve, CD4mem: CD4 memory, CD4nv: CD4 naïve, CD8mem: CD8 memory, CD8nv: CD8 naïve, CRP: C-reactive protein, EGF: Epidermal growth factor, Eos: Eosinophils, Eotaxin: Eosinophil chemotactic protein, FGF: Fibroblast growth factor, G-CSF: Granulocyte colony-stimulating factor, HGF: Hepatocyte growth factor, IFN : Interferon-alpha, IFN gamma: Interferon-gamma, IL: Interleukin, IP10: Interferon gamma-induced protein 10, MCP1: Monocyte chemoattractant protein-1, MIG: Monokine induced by gamma interferon, MIP: Macrophage inflammatory protein, MIP1beta: Macrophage inflammatory protein-1 beta, Mono: Monocytes, Neu: Neutrophils, NK: Natural Killers, Prot: Proteins, Serpine1/PAI-1: Plasminogen activator inhibitor-1, TNF: Tumor necrosis factor-alpha. Treg: T regulatory, WBC: white blood cell.

#### eFigure 4 - Results of the sex-stratified analysis: description of the main signatures

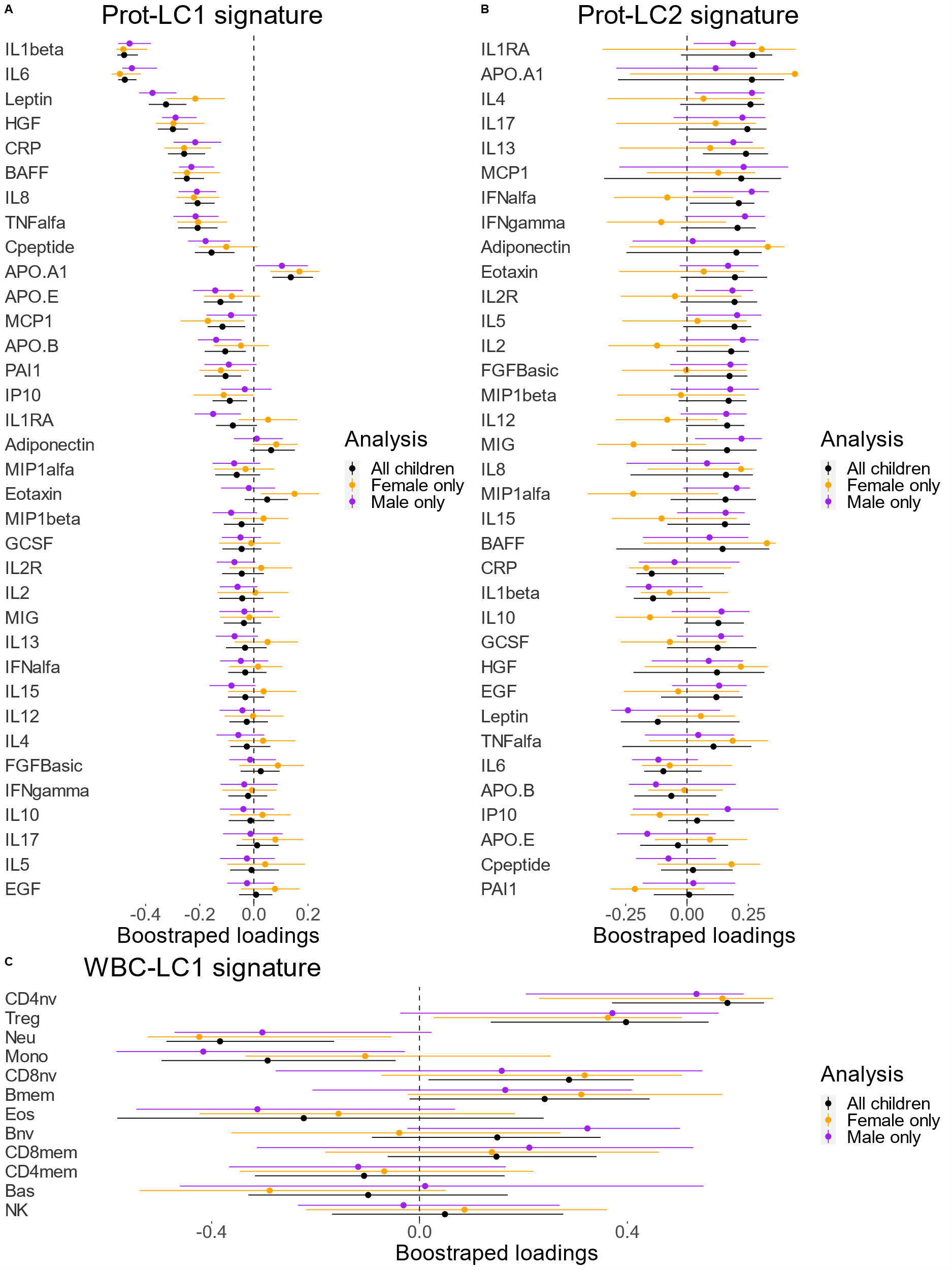

This figure describes biomarker loadings for the main signatures in sex-stratified analyses (see eMethod 5). The point represents biomarkers loadings, i.e. the correlation between each biomarker and a given component, and 95% confidence intervals were computed using bootstraping.

ApoA1: Apolipoprotein A1, ApoB: Apolipoprotein B, ApoE: Apolipoprotein E, BAFF: B-cell activating factor, Bas: Basophils, Bmem: B memory, Bnv: B naïve, CD4mem: CD4 memory, CD4nv: CD4 naïve, CD8mem: CD8 memory, CD8nv: CD8 naïve, CRP: C-reactive protein, EGF: Epidermal growth factor, Eos: Eosinophils, Eotaxin: Eosinophil chemotactic protein, FGF: Fibroblast growth factor, G-CSF: Granulocyte colony-stimulating factor, HGF: Hepatocyte growth factor, IFN : Interferon-alpha, IFN gamma: Interferon-gamma, IL: Interleukin, IP10: Interferon gamma-induced protein 10, MCP1: Monocyte chemoattractant protein-1, MIG: Monokine induced by gamma interferon, MIP: Macrophage inflammatory protein, MIP1beta: Macrophage inflammatory protein-1 beta, Mono: Monocytes, Neu: Neutrophils, NK: Natural Killers, Prot: Proteins, Serpine1/PAI-1: Plasminogen activator inhibitor-1, TNF: Tumor necrosis factor-alpha. Treg: T regulatory, WBC: white blood cell.

#### eFigure 5 – Results of the sensitivity analysis: description of the main signatures

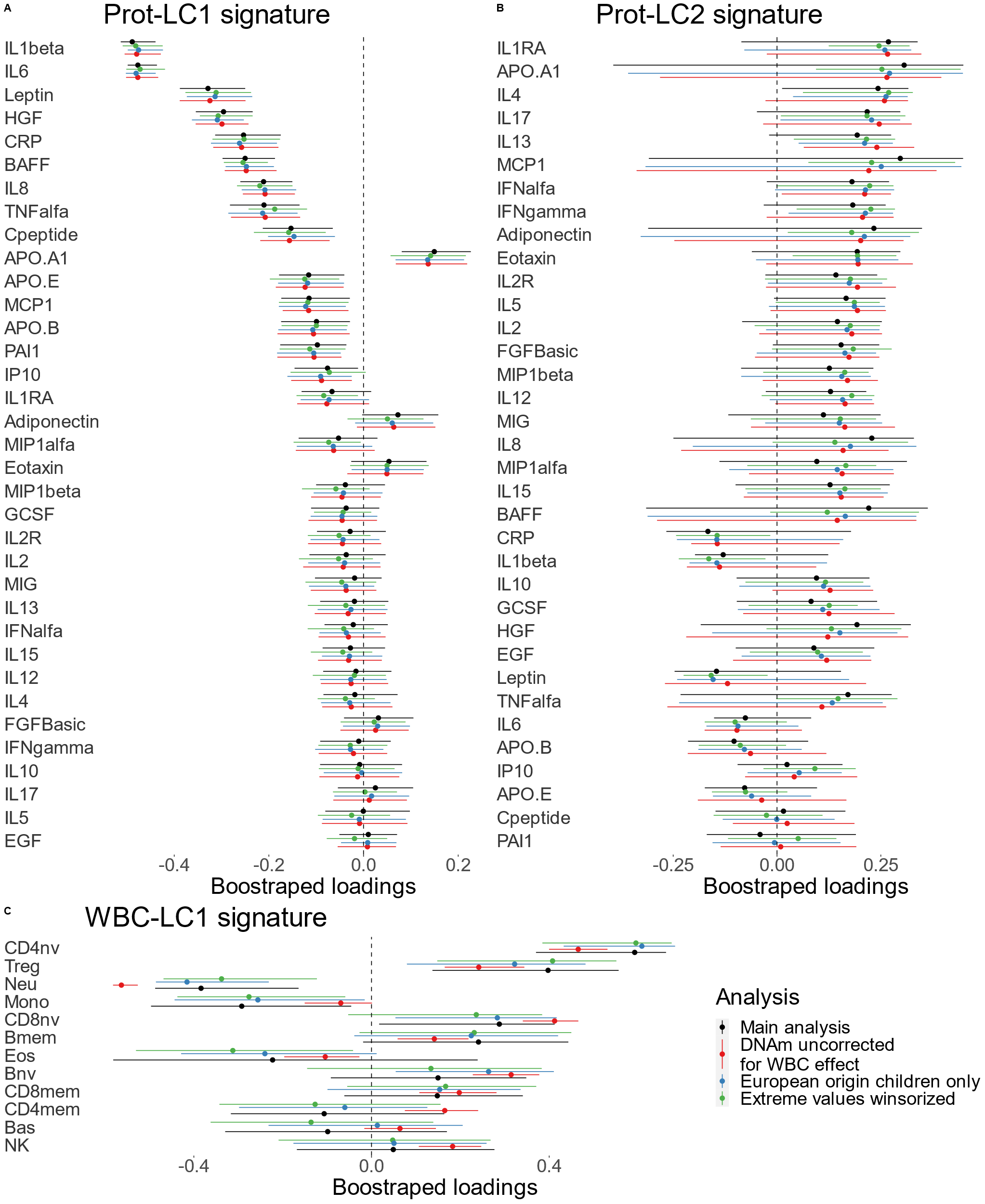
This figure describes biomarker loadings for the main signatures (Prot-LC1, Prot-LC2, WBC-LC1) in the main and sensitivity analyses, at the exception of the leave-one-cohort out analysis (see eMethod 5). The point represents biomarkers loadings, i.e. the correlation between each biomarker and a given component, and 95% confidence intervals were computed using bootstraping.

ApoA1: Apolipoprotein A1, ApoB: Apolipoprotein B, ApoE: Apolipoprotein E, BAFF: B-cell activating factor, Bas: Basophils, Bmem: B memory, Bnv: B naïve, CD4mem: CD4 memory, CD4nv: CD4 naïve, CD8mem: CD8 memory, CD8nv: CD8 naïve, CRP: C-reactive protein, EGF: Epidermal growth factor, Eos: Eosinophils, Eotaxin: Eosinophil chemotactic protein, FGF: Fibroblast growth factor, G-CSF: Granulocyte colony-stimulating factor, HGF: Hepatocyte growth factor, IFN : Interferon-alpha, IFN gamma: Interferon-gamma, IL: Interleukin, IP10: Interferon gamma-induced protein 10, MCP1: Monocyte chemoattractant protein-1, MIG: Monokine induced by gamma interferon, MIP: Macrophage inflammatory protein, MIP1beta: Macrophage inflammatory protein-1 beta, Mono: Monocytes, Neu: Neutrophils, NK: Natural Killers, Prot: Proteins, Serpine1/PAI-1: Plasminogen activator inhibitor-1, TNF: Tumor necrosis factor-alpha. Treg: T regulatory, WBC: white blood cell.

#### eFigure 6 – Results of the leave-one-cohort-out analysis: description of the main signatures

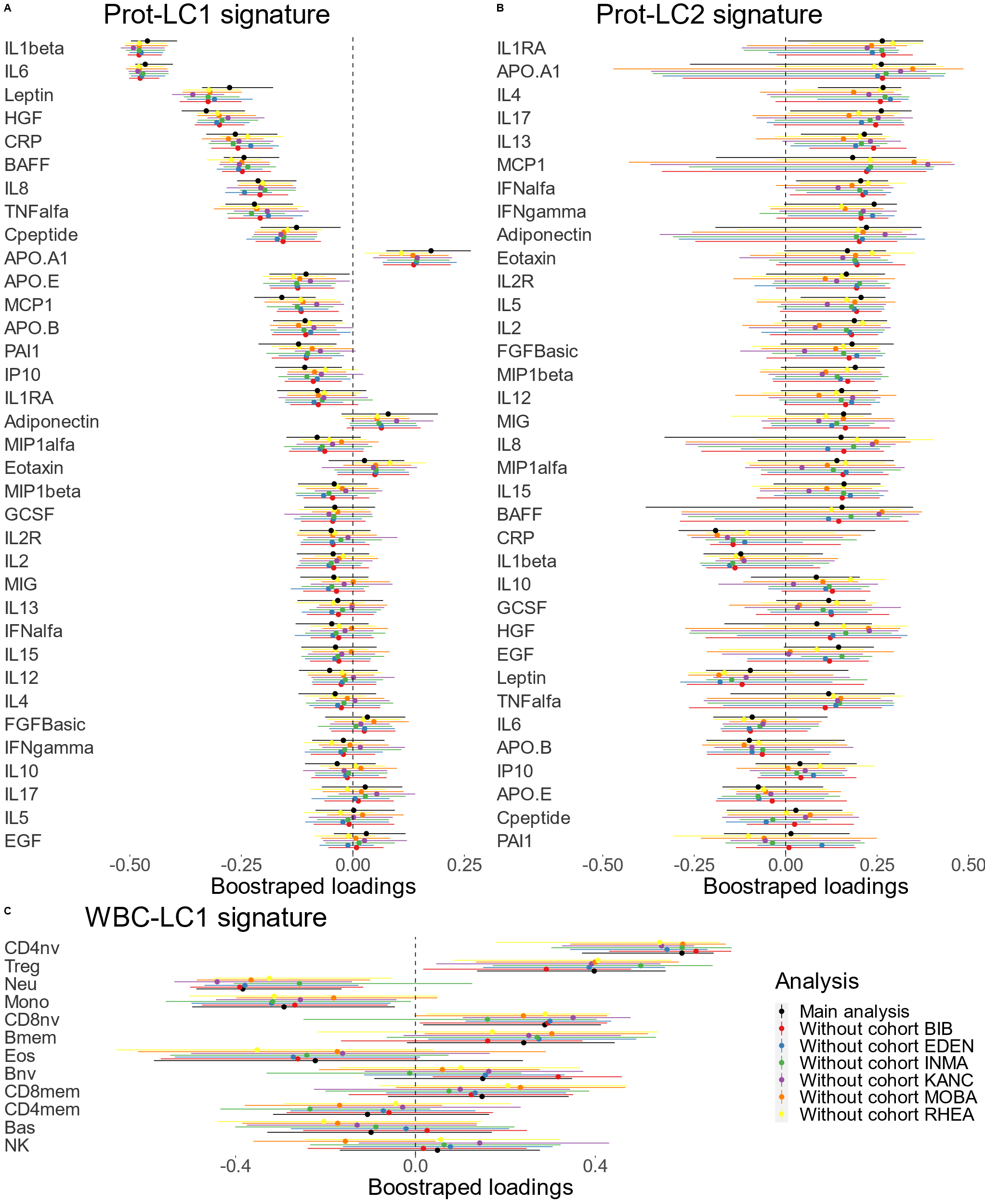
This figure describes biomarker loadings for the main signatures (Prot-LC1, Prot-LC2, WBC-LC1) in the leave-one-cohort out analysis (see eMethod 5). The point represents biomarkers loadings, i.e. the correlation between each biomarker and a given component, and 95% confidence intervals were computed using bootstraping.

ApoA1: Apolipoprotein A1, ApoB: Apolipoprotein B, ApoE: Apolipoprotein E, BAFF: B-cell activating factor, Bas: Basophils, Bmem: B memory, Bnv: B naïve, CD4mem: CD4 memory, CD4nv: CD4 naïve, CD8mem: CD8 memory, CD8nv: CD8 naïve, CRP: C-reactive protein, EGF: Epidermal growth factor, Eos: Eosinophils, Eotaxin: Eosinophil chemotactic protein, FGF: Fibroblast growth factor, G-CSF: Granulocyte colony-stimulating factor, HGF: Hepatocyte growth factor, IFN : Interferon-alpha, IFN gamma: Interferon-gamma, IL: Interleukin, IP10: Interferon gamma-induced protein 10, MCP1: Monocyte chemoattractant protein-1, MIG: Monokine induced by gamma interferon, MIP: Macrophage inflammatory protein, MIP1beta: Macrophage inflammatory protein-1 beta, Mono: Monocytes, Neu: Neutrophils, NK: Natural Killers, Prot: Proteins, Serpine1/PAI-1: Plasminogen activator inhibitor-1, TNF: Tumor necrosis factor-alpha. Treg: T regulatory, WBC: white blood cell.

### Supplementary tables

#### eTable 1 - Transformation applied to continuous postnatal exposures and IQR

| Exposure | Transformation | IQR |
| --- | --- | --- |
| NO2 (preg) - prenatal | Log | 0.3435299 |
| PM10 (preg) - prenatal | None | 11.2685738 |
| PM2.5 (preg) - prenatal | None | 3.8045988 |
| PMabsorbance (preg) - prenatal | Log | 0.3768162 |
| Pressure (t1) - prenatal | None | 21.5352783 |
| Temperature (t1) - prenatal | None | 5.6699142 |
| Humidity (t1) - prenatal | None | 15.0879364 |
| NDVI (100 m) - prenatal | None | 0.2707543 |
| Population density - prenatal | Sqrt | 45.9248811 |
| Built density (300m) - prenatal | Sqrt | 177.8482855 |
| Connectivity (300m) - prenatal | Sqrt | 6.8226691 |
| Accessibility (bus stops 300m) - prenatal | Log | 1.0986169 |
| Facility richness (300m) - prenatal | None | 0.1228070 |
| Facility density (300m) - prenatal | Log | 1.5040811 |
| Land use (300m) - prenatal | None | 0.1758983 |
| Walkability - prenatal | None | 0.1250000 |
| Road traffic load (100 m) - prenatal | Power 1/3 | 119.4879851 |
| Traffic density on nearest road - prenatal | Power 1/3 | 7.9718450 |
| Inverse distance to nearest road - prenatal | Log | 1.1606141 |
| Water THMs - prenatal | Log | 1.7370181 |
| Water Brominated THMs - prenatal | Log | 2.7201134 |
| Water Chloroform - prenatal | Log | 2.5501345 |
| NO2 (year) - postnatal | Log | 0.5869999 |
| PM10 (year) - postnatal | None | 10.9300745 |
| PM2.5 (year) - postnatal | None | 3.1306005 |
| PMabsorbance (year) - postnatal | Log | 0.1934003 |
| Temperature (month) - postnatal | None | 9.3618450 |
| Humidity (month) - postnatal | None | 15.9059140 |
| UV - Vit.D (month) - postnatal | None | 2.0383333 |
| NDVI (100 m) - postnatal | None | 0.2627866 |
| Population density - postnatal | Sqrt | 51.7016005 |
| Built density (300m) - postnatal | Sqrt | 183.5247882 |
| Connectivity density (300 m) - postnatal | Log | 0.7308877 |
| Accessibility (bus stops 300m) - postnatal | Log | 1.2527699 |
| Facility richness (300m) - postnatal | None | 0.1052632 |
| Facility density (300m) - postnatal | Log | 1.4663415 |
| Land use (300m) - postnatal | None | 0.1764468 |
| Walkability index - postnatal | None | 0.1500000 |
| Road traffic load (100 m) - postnatal | Power 1/3 | 120.4592752 |
| Inverse distance to nearest road - postnatal | Log | 1.4447436 |
| Traffic density on nearest road (home) - postnatal | Power 1/3 | 13.6698623 |
| Indoor BTEX - postnatal | Log | 0.4388396 |
| Indoor NO2 - postnatal | Log | 1.2709767 |
| Indoor benzene - postnatal | Log | 0.3086627 |
| Indoor PM2.5 - postnatal | Log | 0.3418614 |
| Indoor PMabsorbance - postnatal | Log | 0.3529038 |
| House crowding - postnatal | Log | 0.4038054 |
| KIDMED score - postnatal | None | 2.0000000 |
| Sleep duration - postnatal | None | 0.9258261 |
| Maternal stress - postnatal | Log | 1.2492030 |
| Moderate and vigorous PA - postnatal | None | 32.7570479 |
| Sedentary behaviour - postnatal | None | 130.7142857 |

*IQR: Interquartile range (Q75%-Q25%), calculated based on the 20 stacked imputed datasets (the original dataset was not included).

Sqrt = square root.

Standardisation on IQR was applied after the transformation (log, power etc) by removing the mean and dividing by the IQR.

#### eTable 2 - Description of the population: health parameters and covariates

| **Variable** | **N = 870** |
| --- | --- |
| **Child health score** | 0.03 (-0.37, 0.46) |
| **Cohort** |  |
| BIB | 134 (15%) |
| EDEN | 112 (13%) |
| INMA | 160 (18%) |
| KANC | 129 (15%) |
| MOBA | 193 (22%) |
| RHEA | 142 (16%) |
| **Child age at follow up** | 8.06 (6.52, 8.89) |
| **Child sex** |  |
| Female | 411 (47%) |
| Male | 459 (53%) |
| **Child ethnicity** |  |
| European | 784 (92%) |
| Non-European | 69 (8.1%) |
| **Maternal education** |  |
| Primary | 121 (14%) |
| Secondary | 301 (35%) |
| Higher | 448 (51%) |
| **Pre-pregnancy**  **Body Mass Index** | 23.8 (21.3, 26.8) |
| **Season of birth** |  |
| Winter | 181 (21%) |
| Spring | 186 (21%) |
| Summer | 244 (28%) |
| Autumn | 259 (30%) |
| **Breastfeeding duration** |  |
| <11 weeks | 273 (31%) |
| 11-35 weeks | 291 (33%) |
| >35 weeks | 306 (35%) |
| **Maternal age at birth** | 31.0 (27.8, 34.0) |

This table gives the description of the health score and covariates in the whole population (n=845). For continuous variables, median and P25, P75 are given. For categorical variables, n (%) is given for each modality.

#### eTable 3 - Description of the population by cohort: health parameters and covariates

| **Variable** | **BIB**,  N = 134 | **EDEN**,  N = 107 | **INMA**,  N = 156 | **KANC**,  N = 125 | **MOBA**,  N = 181 | **RHEA**,  N = 142 | **p-value** |
| --- | --- | --- | --- | --- | --- | --- | --- |
| **Child health score** | -0.21  (-0.59, 0.22) | -0.21  (-0.55, 0.05) | 0.01  (-0.42, 0.33) | 0.29  (-0.14, 0.62) | 0.42  (0.07, 0.73) | -0.13  (-0.46, 0.26) | <0.001 |
| **Child age at follow up** | 6.61  (6.45, 6.79) | 10.83  (10.31, 11.14) | 8.82  (8.42, 9.27) | 6.48  (6.16, 6.85) | 8.55  (8.20, 8.83) | 6.47  (6.36, 6.68) | <0.001 |
| **Child sex** |  |  |  |  |  |  | 0.9 |
| Female | 68 (51%) | 50 (47%) | 68 (44%) | 61 (49%) | 86 (48%) | 64 (45%) |  |
| male | 66 (49%) | 57 (53%) | 88 (56%) | 64 (51%) | 95 (52%) | 78 (55%) |  |
| **Child ethnicity** |  |  |  |  |  |  | <0.001 |
| European origin | 58 (48%) | 107 (100%) | 156 (100%) | 125 (100%) | 172 (97%) | 142 (100%) |  |
| Non-European origin | 62 (52%) | 0 (0%) | 0 (0%) | 0 (0%) | 6 (3.4%) | 0 (0%) |  |
| **Maternal education** |  |  |  |  |  |  | <0.001 |
| Primary | 63 (47%) | 8 (7.5%) | 37 (24%) | 8 (6.4%) | 0 (0%) | 4 (2.8%) |  |
| Secondary | 23 (17%) | 41 (38%) | 68 (44%) | 43 (34%) | 35 (19%) | 80 (56%) |  |
| Higher | 48 (36%) | 58 (54%) | 51 (33%) | 74 (59%) | 146 (81%) | 58 (41%) |  |
| **Pre-pregnancy Body Mass Index** | 26.8  (24.4, 31.3) | 22.0  (20.1, 25.7) | 23.0  (21.1, 25.5) | 26.7  (24.6, 30.5) | 22.1  (20.6, 24.0) | 23.3  (21.5, 25.1) | <0.001 |
| **Season of birth** |  |  |  |  |  |  | <0.001 |
| Winter | 10 (7.5%) | 26 (24%) | 28 (18%) | 23 (18%) | 44 (24%) | 45 (32%) |  |
| Spring | 22 (16%) | 16 (15%) | 47 (30%) | 44 (35%) | 24 (13%) | 27 (19%) |  |
| Summer | 51 (38%) | 33 (31%) | 50 (32%) | 28 (22%) | 51 (28%) | 25 (18%) |  |
| Autumn | 51 (38%) | 32 (30%) | 31 (20%) | 30 (24%) | 62 (34%) | 45 (32%) |  |
| **Breastfeeding duration** |  |  |  |  |  |  | <0.001 |
| <11 weeks | 42 (31%) | 64 (60%) | 48 (31%) | 25 (20%) | 27 (15%) | 63 (44%) |  |
| 11-35 weeks | 57 (43%) | 36 (34%) | 66 (42%) | 37 (30%) | 34 (19%) | 53 (37%) |  |
| >35 weeks | 35 (26%) | 7 (6.5%) | 42 (27%) | 63 (50%) | 120 (66%) | 26 (18%) |  |
| **Maternal age at birth** | 28.0  (25.0, 32.0) | 30.3  (28.2, 34.1) | 32.1  (29.6, 34.7) | 29.0  (25.6, 32.4) | 33.0  (30.0, 35.0) | 30.9  (27.2, 33.4) | <0.001 |

This table gives the description of the health score and covariates in each cohort (total n=845). For continuous variables, median and P25, P75 are given. For categorical variables, n (%) is given for each modality.

#### eTable 4 - Correlations between signatures estimated by the multiblock model

|  | Prot-LC1 | Prot-LC2 | Prot-LC3 | WBC-LC1 | WBC-LC2 | WBC-LC3 | DNAm-LC1 |
| --- | --- | --- | --- | --- | --- | --- | --- |
| Prot-LC1 | 1.00 | 0.00 | 0.00 | 0.21 | -0.01 | 0.02 | 0.06 |
| Prot-LC2 | 0.00 | 1.00 | 0.00 | -0.05 | 0.00 | 0.01 | -0.01 |
| Prot-LC3 | 0.00 | 0.00 | 1.00 | 0.01 | 0.01 | 0.11 | 0.07 |
| WBC-LC1 | 0.21 | -0.05 | 0.01 | 1.00 | 0.00 | 0.00 | 0.00 |
| WBC-LC2 | -0.01 | 0.00 | 0.01 | 0.00 | 1.00 | 0.00 | 0.04 |
| WBC-LC3 | 0.02 | 0.01 | 0.11 | 0.00 | 0.00 | 1.00 | 0.16 |
| DNAm-LC1 | 0.06 | -0.01 | 0.07 | 0.00 | 0.04 | 0.16 | 1.00 |

This table gives the pearson correlation coefficients between signatures in the whole database (n=845).

#### eTable 5 – Significant associations between the immune signatures and the health scores

| Signatures | Health score | beta (CI 95%) | p |
| --- | --- | --- | --- |
| WBC-LC1 | Multi Domain Health score | 0.24 [0.15; 0.33] | 4.0e-07 |
| WBC-LC2 | Multi Domain Health score | 0.21 [0.096; 0.33] | 3.5e-04 |
| Prot-LC1 | Multi Domain Health score | 0.74 [0.64; 0.84] | 0.0e+00 |
| Prot-LC2 | Multi Domain Health score | 0.17 [0.085; 0.26] | 1.2e-04 |
| Prot-LC3 | Multi Domain Health score | 0.36 [0.21; 0.52] | 5.9e-06 |
| DNAm-LC1 | Multi Domain Health score | 12.0 [6.4; 17.0] | 1.3e-05 |
| WBC-LC2 | Respiratory/allergic subscore | 0.25 [0.059; 0.44] | 1.1e-02 |
| Prot-LC1 | Respiratory/allergic subscore | 0.31 [0.12; 0.49] | 1.4e-03 |
| DNAm-LC1 | Respiratory/allergic subscore | 19 [10; 27] | 1.9e-05 |
| WBC-LC1 | Cardiometabolic subscore | 0.26 [0.093; 0.42] | 2.1e-03 |
| WBC-LC3 | Cardiometabolic subscore | 0.2 [0.005; 0.4] | 4.5e-02 |
| Prot-LC1 | Cardiometabolic subscore | 1.7 [1.5; 1.9] | 0.0e+00 |
| Prot-LC2 | Cardiometabolic subscore | 0.49 [0.34; 0.64] | 0.0e+00 |
| Prot-LC3 | Cardiometabolic subscore | 0.84 [0.57; 1.1] | 0.0e+00 |
| DNAm-LC1 | Cardiometabolic subscore | 14 [4.4; 23] | 3.9e-03 |
| WBC-LC1 | Neurodevelopmental subscore | 0.31 [0.16; 0.46] | 5.4e-05 |
| WBC-LC2 | Neurodevelopmental subscore | 0.25 [0.064; 0.44] | 8.9e-03 |
| WBC-LC3 | Neurodevelopmental subscore | -0.22 [-0.4; -0.037] | 1.9e-02 |
| Prot-LC1 | Neurodevelopmental subscore | 0.23 [0.044; 0.41] | 1.5e-02 |

This table gives the estimations for the significant associations between signatures (Component) and health scores, with there 95% confidence interval (CI), p-values (p).

#### eTable 6 - Significant associations between exposures and the signatures

| **Exposure** | **Modality** | **Signature** | **beta (CI 95%)** | **p** |
| --- | --- | --- | --- | --- |
| Accessibility (bus lines 300m) - postnatal |  | Prot-LC1 | 0.03 [-0.02; 0.08] | 0,2 |
| Accessibility (bus lines 300m) - prenatal |  | Prot-LC1 | 0.05 [0; 0.1] | 0,03 |
| Accessibility (bus stops 300m) - postnatal |  | Prot-LC1 | 0 [-0.04; 0.05] | 0,87 |
| Accessibility (bus stops 300m) - prenatal |  | Prot-LC1 | 0.01 [-0.02; 0.04] | 0,62 |
| Bakery products intake - postnatal | High vs low | Prot-LC1 | 0.03 [-0.03; 0.09] | 0,31 |
| Bakery products intake - postnatal | Medium vs low | Prot-LC1 | -0.01 [-0.07; 0.04] | 0,55 |
| Blue spaces (300 m) - postnatal | Yes | Prot-LC1 | 0.09 [0; 0.17] | 0,04 |
| Blue spaces (300 m) - prenatal | Yes | Prot-LC1 | 0.08 [-0.01; 0.16] | 0,09 |
| Bread intake - postnatal | Medium vs low | Prot-LC1 | -0.04 [-0.09; 0.01] | 0,16 |
| Bread intake - postnatal | High vs low | Prot-LC1 | -0.01 [-0.07; 0.04] | 0,58 |
| Breakfast cereals intake - postnatal | High vs low | Prot-LC1 | 0.07 [0.01; 0.12] | 0,01 |
| Breakfast cereals intake - postnatal | Medium vs low | Prot-LC1 | 0.02 [-0.03; 0.07] | 0,43 |
| Built density (300m) - postnatal |  | Prot-LC1 | -0.01 [-0.04; 0.01] | 0,39 |
| Built density (300m) - prenatal |  | Prot-LC1 | -0.01 [-0.04; 0.02] | 0,5 |
| Caffeinated drinks - postnatal | High vs low | Prot-LC1 | -0.03 [-0.08; 0.03] | 0,34 |
| Caffeinated drinks - postnatal | Medium vs low | Prot-LC1 | 0.02 [-0.05; 0.08] | 0,62 |
| Cat at home - postnatal | Yes | Prot-LC1 | -0.01 [-0.07; 0.04] | 0,64 |
| Cereals intake - postnatal | Medium vs low | Prot-LC1 | 0.01 [-0.05; 0.06] | 0,8 |
| Cereals intake - postnatal | High vs low | Prot-LC1 | 0.01 [-0.05; 0.06] | 0,8 |
| Cereals intake - prenatal | Medium vs low | Prot-LC1 | -0.03 [-0.08; 0.02] | 0,23 |
| Cereals intake - prenatal | High vs low | Prot-LC1 | -0.02 [-0.07; 0.04] | 0,54 |
| Connectivity (300m) - prenatal |  | Prot-LC1 | 0.01 [-0.02; 0.04] | 0,47 |
| Connectivity density (300 m) - postnatal |  | Prot-LC1 | 0 [-0.03; 0.04] | 0,82 |
| Contact with family and friends - postnatal |  | Prot-LC1 | -0.03 [-0.08; 0.02] | 0,23 |
| Contact with family and friends - postnatal |  | Prot-LC1 | -0.01 [-0.11; 0.09] | 0,81 |
| Dairy products intake - postnatal | Medium vs low | Prot-LC1 | -0.05 [-0.1; 0] | 0,06 |
| Dairy products intake - postnatal | High vs low | Prot-LC1 | -0.04 [-0.1; 0.01] | 0,14 |
| Dog at home - postnatal | Yes | Prot-LC1 | 0.01 [-0.05; 0.07] | 0,79 |
| Exposure to smoke - postnatal |  | Prot-LC1 | -0.03 [-0.07; 0.01] | 0,2 |
| Facility density (300m) - postnatal |  | Prot-LC1 | -0.01 [-0.05; 0.03] | 0,66 |
| Facility density (300m) - prenatal |  | Prot-LC1 | 0.01 [-0.02; 0.05] | 0,5 |
| Facility richness (300m) - postnatal |  | Prot-LC1 | 0 [-0.04; 0.03] | 0,79 |
| Facility richness (300m) - prenatal |  | Prot-LC1 | 0 [-0.03; 0.04] | 0,8 |
| Family affluence score - postnatal |  | Prot-LC1 | 0.01 [-0.06; 0.09] | 0,77 |
| Family affluence score - postnatal |  | Prot-LC1 | 0 [-0.07; 0.08] | 0,94 |
| Family size - postnatal |  | Prot-LC1 | 0.04 [0.01; 0.06] | 0,01 |
| Fastfood intake - postnatal | High vs low | Prot-LC1 | -0.01 [-0.08; 0.05] | 0,66 |
| Fastfood intake - postnatal | Medium vs low | Prot-LC1 | 0 [-0.05; 0.05] | 0,94 |
| Fish and seafood intake - postnatal | High vs low | Prot-LC1 | 0.04 [-0.02; 0.09] | 0,19 |
| Fish and seafood intake - postnatal | Medium vs low | Prot-LC1 | 0.03 [-0.02; 0.09] | 0,22 |
| Fish and seafood intake - prenatal | High vs low | Prot-LC1 | 0.01 [-0.04; 0.06] | 0,65 |
| Fish and seafood intake - prenatal | Medium vs low | Prot-LC1 | 0 [-0.05; 0.06] | 0,92 |
| Fruits intake - postnatal | High vs low | Prot-LC1 | 0.02 [-0.04; 0.07] | 0,5 |
| Fruits intake - postnatal | Medium vs low | Prot-LC1 | -0.01 [-0.06; 0.05] | 0,82 |
| Green spaces (300 m) - postnatal | Yes | Prot-LC1 | 0.04 [-0.01; 0.1] | 0,13 |
| Green spaces (300 m) - prenatal | Yes | Prot-LC1 | 0.02 [-0.03; 0.07] | 0,4 |
| Humidity (month) - postnatal |  | Prot-LC1 | -0.01 [-0.04; 0.02] | 0,48 |
| Humidity (t1) - prenatal |  | Prot-LC1 | 0 [-0.04; 0.03] | 0,94 |
| Indoor BTEX - postnatal |  | Prot-LC1 | -0.01 [-0.04; 0.02] | 0,4 |
| Indoor NO2 - postnatal |  | Prot-LC1 | -0.01 [-0.04; 0.03] | 0,73 |
| Indoor PM2.5 - postnatal |  | Prot-LC1 | -0.03 [-0.05; 0] | 0,02 |
| Indoor PMabsorbance - postnatal |  | Prot-LC1 | -0.03 [-0.05; -0.01] | 0 |
| Indoor benzene - postnatal |  | Prot-LC1 | -0.03 [-0.06; -0.01] | 0,01 |
| Inverse distance to nearest road - postnatal |  | Prot-LC1 | 0 [-0.03; 0.03] | 0,98 |
| Inverse distance to nearest road - prenatal |  | Prot-LC1 | 0.02 [-0.01; 0.04] | 0,15 |
| KIDMED score - postnatal |  | Prot-LC1 | 0.01 [-0.01; 0.04] | 0,3 |
| Land use (300m) - postnatal |  | Prot-LC1 | 0.03 [0; 0.06] | 0,07 |
| Land use (300m) - prenatal |  | Prot-LC1 | 0.02 [0; 0.05] | 0,1 |
| Maternal smoking (active and ETS) - prenatal |  | Prot-LC1 | -0.04 [-0.08; 0.01] | 0,12 |
| Meat intake - postnatal | High vs low | Prot-LC1 | -0.03 [-0.08; 0.03] | 0,32 |
| Meat intake - postnatal | Medium vs low | Prot-LC1 | -0.01 [-0.07; 0.04] | 0,66 |
| Meat intake - prenatal | Medium vs low | Prot-LC1 | -0.03 [-0.09; 0.02] | 0,22 |
| Meat intake - prenatal | High vs low | Prot-LC1 | 0.01 [-0.04; 0.07] | 0,66 |
| Moderate and vigorous PA - postnatal |  | Prot-LC1 | 0.01 [-0.02; 0.04] | 0,65 |
| NDVI (100 m) - postnatal |  | Prot-LC1 | 0.01 [-0.03; 0.04] | 0,61 |
| NDVI (100 m) - prenatal |  | Prot-LC1 | 0.01 [-0.03; 0.04] | 0,78 |
| Organic food intake - postnatal | High vs low | Prot-LC1 | 0.07 [0.01; 0.13] | 0,01 |
| Organic food intake - postnatal | Medium vs low | Prot-LC1 | 0.03 [-0.03; 0.07] | 0,34 |
| Other pets at home - postnatal |  | Prot-LC1 | 0.01 [-0.03; 0.06] | 0,56 |
| Outdoor NO2 (preg) - prenatal |  | Prot-LC1 | 0 [-0.03; 0.02] | 0,79 |
| Outdoor NO2 (year) - postnatal |  | Prot-LC1 | -0.03 [-0.07; 0.01] | 0,11 |
| Outdoor PM10 (preg) - prenatal |  | Prot-LC1 | 0 [-0.03; 0.03] | 0,88 |
| Outdoor PM10 (year) - postnatal |  | Prot-LC1 | -0.01 [-0.04; 0.02] | 0,42 |
| Outdoor PM2.5 (preg) - prenatal |  | Prot-LC1 | 0 [-0.03; 0.03] | 0,91 |
| Outdoor PM2.5 (year) - postnatal |  | Prot-LC1 | -0.01 [-0.03; 0.01] | 0,26 |
| Outdoor PMabsorbance (preg) - prenatal |  | Prot-LC1 | -0.01 [-0.04; 0.03] | 0,67 |
| Outdoor PMabsorbance (year) - postnatal |  | Prot-LC1 | -0.02 [-0.05; 0] | 0,11 |
| Parental smoking - postnatal |  | Prot-LC1 | -0.05 [-0.13; 0.02] | 0,15 |
| Parental smoking - postnatal |  | Prot-LC1 | -0.03 [-0.08; 0.02] | 0,26 |
| Population density - postnatal |  | Prot-LC1 | -0.01 [-0.04; 0.01] | 0,27 |
| Population density - prenatal |  | Prot-LC1 | 0 [-0.03; 0.02] | 0,71 |
| Potatoes intake - postnatal | Medium vs low | Prot-LC1 | -0.03 [-0.08; 0.01] | 0,18 |
| Potatoes intake - postnatal | High vs low | Prot-LC1 | -0.04 [-0.1; 0.03] | 0,27 |
| Pressure (t1) - prenatal |  | Prot-LC1 | 0 [-0.04; 0.03] | 0,88 |
| Processed meat intake - postnatal | High vs low | Prot-LC1 | 0.03 [-0.03; 0.09] | 0,35 |
| Processed meat intake - postnatal | Medium vs low | Prot-LC1 | 0.02 [-0.03; 0.07] | 0,53 |
| Ready made food intake - postnatal | High vs low | Prot-LC1 | -0.02 [-0.07; 0.03] | 0,49 |
| Ready made food intake - postnatal | Medium vs low | Prot-LC1 | -0.02 [-0.07; 0.04] | 0,51 |
| Road traffic load (100 m) - postnatal |  | Prot-LC1 | -0.03 [-0.07; 0.01] | 0,12 |
| Road traffic load (100 m) - prenatal |  | Prot-LC1 | -0.01 [-0.05; 0.03] | 0,5 |
| Sedentary behaviour - postnatal |  | Prot-LC1 | 0.01 [-0.01; 0.03] | 0,33 |
| Sleep duration - postnatal |  | Prot-LC1 | 0.02 [-0.01; 0.05] | 0,26 |
| Social participation - postnatal |  | Prot-LC1 | 0.05 [0; 0.1] | 0,03 |
| Social participation - postnatal |  | Prot-LC1 | -0.04 [-0.1; 0.03] | 0,24 |
| Soda intake - postnatal | Medium vs low | Prot-LC1 | 0.03 [-0.02; 0.08] | 0,31 |
| Soda intake - postnatal | High vs low | Prot-LC1 | -0.01 [-0.06; 0.05] | 0,83 |
| Sweets intake - postnatal | Medium vs low | Prot-LC1 | 0.04 [-0.01; 0.1] | 0,13 |
| Sweets intake - postnatal | High vs low | Prot-LC1 | 0.01 [-0.04; 0.07] | 0,57 |
| Temperature (previous month) - postnatal |  | Prot-LC1 | 0.01 [-0.01; 0.04] | 0,33 |
| Temperature (t1) - prenatal |  | Prot-LC1 | 0.01 [-0.02; 0.04] | 0,45 |
| Total fat intake - postnatal | High vs low | Prot-LC1 | 0.02 [-0.03; 0.07] | 0,46 |
| Total fat intake - postnatal | Medium vs low | Prot-LC1 | 0.01 [-0.04; 0.07] | 0,64 |
| Traffic density on major road (home) - postnatal | Yes | Prot-LC1 | -0.01 [-0.07; 0.04] | 0,61 |
| Traffic density on major road - prenatal | Yes | Prot-LC1 | -0.04 [-0.09; 0] | 0,06 |
| Traffic density on nearest road (home) - postnatal |  | Prot-LC1 | -0.02 [-0.05; 0.02] | 0,3 |
| Traffic density on nearest road - prenatal |  | Prot-LC1 | -0.01 [-0.03; 0.02] | 0,58 |
| UV - Vit.D (month) - postnatal |  | Prot-LC1 | 0.02 [-0.01; 0.05] | 0,24 |
| Vegetables intake - postnatal | High vs low | Prot-LC1 | 0.06 [0.01; 0.11] | 0,02 |
| Vegetables intake - postnatal | Medium vs low | Prot-LC1 | -0.04 [-0.1; 0.02] | 0,18 |
| Walkability - prenatal |  | Prot-LC1 | 0.03 [0; 0.07] | 0,07 |
| Walkability index - postnatal |  | Prot-LC1 | 0.01 [-0.02; 0.05] | 0,49 |
| Water Brominated THMs - prenatal |  | Prot-LC1 | -0.01 [-0.04; 0.02] | 0,52 |
| Water Chloroform - prenatal |  | Prot-LC1 | 0 [-0.04; 0.04] | 0,9 |
| Water THMs - prenatal |  | Prot-LC1 | 0 [-0.03; 0.04] | 0,85 |
| Yogurt intake - postnatal | High vs low | Prot-LC1 | 0.04 [-0.01; 0.1] | 0,08 |
| Yogurt intake - postnatal | Medium vs low | Prot-LC1 | 0.01 [-0.05; 0.06] | 0,78 |
| Accessibility (bus lines 300m) - postnatal |  | Prot-LC2 | -0.02 [-0.08; 0.04] | 0,6 |
| Accessibility (bus lines 300m) - prenatal |  | Prot-LC2 | 0.01 [-0.05; 0.07] | 0,81 |
| Accessibility (bus stops 300m) - postnatal |  | Prot-LC2 | 0 [-0.06; 0.06] | 0,95 |
| Accessibility (bus stops 300m) - prenatal |  | Prot-LC2 | 0 [-0.04; 0.04] | 0,97 |
| Bakery products intake - postnatal | Medium vs low | Prot-LC2 | 0.08 [0.01; 0.15] | 0,02 |
| Bakery products intake - postnatal | High vs low | Prot-LC2 | 0.06 [-0.01; 0.13] | 0,1 |
| Blue spaces (300 m) - postnatal | Yes | Prot-LC2 | 0.03 [-0.07; 0.14] | 0,56 |
| Blue spaces (300 m) - prenatal | Yes | Prot-LC2 | 0.05 [-0.06; 0.16] | 0,4 |
| Bread intake - postnatal | Medium vs low | Prot-LC2 | -0.04 [-0.1; 0.03] | 0,26 |
| Bread intake - postnatal | High vs low | Prot-LC2 | -0.01 [-0.08; 0.06] | 0,82 |
| Breakfast cereals intake - postnatal | High vs low | Prot-LC2 | -0.02 [-0.09; 0.05] | 0,6 |
| Breakfast cereals intake - postnatal | Medium vs low | Prot-LC2 | -0.01 [-0.08; 0.06] | 0,79 |
| Built density (300m) - postnatal |  | Prot-LC2 | -0.01 [-0.04; 0.02] | 0,49 |
| Built density (300m) - prenatal |  | Prot-LC2 | -0.01 [-0.04; 0.03] | 0,71 |
| Caffeinated drinks - postnatal | High vs low | Prot-LC2 | -0.04 [-0.11; 0.03] | 0,21 |
| Caffeinated drinks - postnatal | Medium vs low | Prot-LC2 | 0.01 [-0.07; 0.09] | 0,82 |
| Cat at home - postnatal | Yes | Prot-LC2 | 0.03 [-0.04; 0.11] | 0,38 |
| Cereals intake - postnatal | Medium vs low | Prot-LC2 | -0.04 [-0.11; 0.03] | 0,31 |
| Cereals intake - postnatal | High vs low | Prot-LC2 | -0.02 [-0.09; 0.05] | 0,63 |
| Cereals intake - prenatal | Medium vs low | Prot-LC2 | -0.03 [-0.09; 0.04] | 0,45 |
| Cereals intake - prenatal | High vs low | Prot-LC2 | -0.02 [-0.09; 0.05] | 0,64 |
| Connectivity (300m) - prenatal |  | Prot-LC2 | 0.01 [-0.03; 0.05] | 0,56 |
| Connectivity density (300 m) - postnatal |  | Prot-LC2 | 0.01 [-0.04; 0.05] | 0,74 |
| Contact with family and friends - postnatal |  | Prot-LC2 | -0.11 [-0.24; 0.02] | 0,09 |
| Contact with family and friends - postnatal |  | Prot-LC2 | -0.05 [-0.12; 0.01] | 0,1 |
| Dairy products intake - postnatal | Medium vs low | Prot-LC2 | 0.06 [-0.01; 0.13] | 0,09 |
| Dairy products intake - postnatal | High vs low | Prot-LC2 | 0.04 [-0.04; 0.1] | 0,34 |
| Dog at home - postnatal | Yes | Prot-LC2 | -0.09 [-0.17; -0.01] | 0,03 |
| Exposure to smoke - postnatal |  | Prot-LC2 | 0.02 [-0.04; 0.08] | 0,52 |
| Facility density (300m) - postnatal |  | Prot-LC2 | -0.02 [-0.07; 0.03] | 0,47 |
| Facility density (300m) - prenatal |  | Prot-LC2 | 0 [-0.04; 0.05] | 0,88 |
| Facility richness (300m) - postnatal |  | Prot-LC2 | -0.02 [-0.06; 0.03] | 0,47 |
| Facility richness (300m) - prenatal |  | Prot-LC2 | 0.01 [-0.04; 0.05] | 0,76 |
| Family affluence score - postnatal |  | Prot-LC2 | 0.07 [-0.03; 0.17] | 0,16 |
| Family affluence score - postnatal |  | Prot-LC2 | 0.06 [-0.04; 0.15] | 0,26 |
| Family size - postnatal |  | Prot-LC2 | -0.01 [-0.05; 0.02] | 0,38 |
| Fastfood intake - postnatal | Medium vs low | Prot-LC2 | 0.03 [-0.03; 0.09] | 0,37 |
| Fastfood intake - postnatal | High vs low | Prot-LC2 | -0.01 [-0.1; 0.08] | 0,88 |
| Fish and seafood intake - postnatal | Medium vs low | Prot-LC2 | -0.02 [-0.09; 0.04] | 0,5 |
| Fish and seafood intake - postnatal | High vs low | Prot-LC2 | -0.01 [-0.07; 0.06] | 0,86 |
| Fish and seafood intake - prenatal | Medium vs low | Prot-LC2 | 0.04 [-0.03; 0.11] | 0,29 |
| Fish and seafood intake - prenatal | High vs low | Prot-LC2 | 0.01 [-0.06; 0.07] | 0,86 |
| Fruits intake - postnatal | Medium vs low | Prot-LC2 | 0.04 [-0.03; 0.11] | 0,26 |
| Fruits intake - postnatal | High vs low | Prot-LC2 | 0.01 [-0.06; 0.08] | 0,72 |
| Green spaces (300 m) - postnatal | Yes | Prot-LC2 | -0.01 [-0.08; 0.06] | 0,72 |
| Green spaces (300 m) - prenatal | Yes | Prot-LC2 | 0.03 [-0.04; 0.09] | 0,42 |
| Humidity (month) - postnatal |  | Prot-LC2 | 0.02 [-0.02; 0.06] | 0,37 |
| Humidity (t1) - prenatal |  | Prot-LC2 | 0 [-0.04; 0.04] | 0,96 |
| Indoor BTEX - postnatal |  | Prot-LC2 | -0.02 [-0.06; 0.02] | 0,29 |
| Indoor NO2 - postnatal |  | Prot-LC2 | -0.02 [-0.07; 0.02] | 0,3 |
| Indoor PM2.5 - postnatal |  | Prot-LC2 | -0.03 [-0.06; 0] | 0,02 |
| Indoor PMabsorbance - postnatal |  | Prot-LC2 | -0.01 [-0.04; 0.02] | 0,54 |
| Indoor benzene - postnatal |  | Prot-LC2 | -0.02 [-0.05; 0.02] | 0,35 |
| Inverse distance to nearest road - postnatal |  | Prot-LC2 | -0.01 [-0.05; 0.03] | 0,65 |
| Inverse distance to nearest road - prenatal |  | Prot-LC2 | 0 [-0.03; 0.04] | 0,78 |
| KIDMED score - postnatal |  | Prot-LC2 | 0 [-0.04; 0.03] | 0,78 |
| Land use (300m) - postnatal |  | Prot-LC2 | 0.01 [-0.02; 0.05] | 0,46 |
| Land use (300m) - prenatal |  | Prot-LC2 | 0.01 [-0.03; 0.04] | 0,65 |
| Maternal smoking (active and ETS) - prenatal |  | Prot-LC2 | -0.02 [-0.08; 0.04] | 0,47 |
| Meat intake - postnatal | Medium vs low | Prot-LC2 | 0 [-0.07; 0.07] | 0,98 |
| Meat intake - postnatal | High vs low | Prot-LC2 | 0 [-0.07; 0.07] | 0,99 |
| Meat intake - prenatal | High vs low | Prot-LC2 | -0.04 [-0.11; 0.03] | 0,25 |
| Meat intake - prenatal | Medium vs low | Prot-LC2 | 0 [-0.07; 0.07] | 0,96 |
| Moderate and vigorous PA - postnatal |  | Prot-LC2 | 0.01 [-0.03; 0.05] | 0,59 |
| NDVI (100 m) - postnatal |  | Prot-LC2 | 0.01 [-0.03; 0.06] | 0,58 |
| NDVI (100 m) - prenatal |  | Prot-LC2 | 0.01 [-0.04; 0.06] | 0,75 |
| Organic food intake - postnatal | High vs low | Prot-LC2 | 0 [-0.07; 0.08] | 0,92 |
| Organic food intake - postnatal | Medium vs low | Prot-LC2 | 0 [-0.06; 0.07] | 0,95 |
| Other pets at home - postnatal |  | Prot-LC2 | -0.03 [-0.09; 0.03] | 0,37 |
| Outdoor NO2 (preg) - prenatal |  | Prot-LC2 | 0 [-0.03; 0.03] | 1 |
| Outdoor NO2 (year) - postnatal |  | Prot-LC2 | -0.01 [-0.06; 0.04] | 0,62 |
| Outdoor PM10 (preg) - prenatal |  | Prot-LC2 | 0 [-0.04; 0.03] | 0,82 |
| Outdoor PM10 (year) - postnatal |  | Prot-LC2 | -0.01 [-0.05; 0.03] | 0,58 |
| Outdoor PM2.5 (preg) - prenatal |  | Prot-LC2 | 0 [-0.04; 0.04] | 0,87 |
| Outdoor PM2.5 (year) - postnatal |  | Prot-LC2 | 0 [-0.02; 0.03] | 0,8 |
| Outdoor PMabsorbance (preg) - prenatal |  | Prot-LC2 | 0.01 [-0.03; 0.05] | 0,7 |
| Outdoor PMabsorbance (year) - postnatal |  | Prot-LC2 | -0.01 [-0.04; 0.03] | 0,67 |
| Parental smoking - postnatal |  | Prot-LC2 | -0.07 [-0.17; 0.02] | 0,13 |
| Parental smoking - postnatal |  | Prot-LC2 | -0.02 [-0.09; 0.04] | 0,54 |
| Population density - postnatal |  | Prot-LC2 | -0.02 [-0.05; 0.01] | 0,3 |
| Population density - prenatal |  | Prot-LC2 | -0.01 [-0.04; 0.02] | 0,6 |
| Potatoes intake - postnatal | Medium vs low | Prot-LC2 | -0.04 [-0.11; 0.02] | 0,2 |
| Potatoes intake - postnatal | High vs low | Prot-LC2 | -0.04 [-0.12; 0.04] | 0,32 |
| Pressure (t1) - prenatal |  | Prot-LC2 | -0.01 [-0.06; 0.03] | 0,62 |
| Processed meat intake - postnatal | Medium vs low | Prot-LC2 | 0.04 [-0.03; 0.1] | 0,27 |
| Processed meat intake - postnatal | High vs low | Prot-LC2 | 0 [-0.07; 0.08] | 0,95 |
| Ready made food intake - postnatal | Medium vs low | Prot-LC2 | 0 [-0.07; 0.07] | 0,98 |
| Ready made food intake - postnatal | High vs low | Prot-LC2 | 0 [-0.07; 0.07] | 0,99 |
| Road traffic load (100 m) - postnatal |  | Prot-LC2 | -0.01 [-0.06; 0.04] | 0,76 |
| Road traffic load (100 m) - prenatal |  | Prot-LC2 | -0.02 [-0.07; 0.03] | 0,37 |
| Sedentary behaviour - postnatal |  | Prot-LC2 | 0 [-0.03; 0.03] | 0,91 |
| Sleep duration - postnatal |  | Prot-LC2 | -0.01 [-0.04; 0.03] | 0,78 |
| Social participation - postnatal |  | Prot-LC2 | -0.03 [-0.09; 0.04] | 0,42 |
| Social participation - postnatal |  | Prot-LC2 | 0.02 [-0.07; 0.1] | 0,69 |
| Soda intake - postnatal | High vs low | Prot-LC2 | -0.03 [-0.1; 0.05] | 0,5 |
| Soda intake - postnatal | Medium vs low | Prot-LC2 | 0.02 [-0.05; 0.08] | 0,57 |
| Sweets intake - postnatal | Medium vs low | Prot-LC2 | -0.05 [-0.12; 0.01] | 0,13 |
| Sweets intake - postnatal | High vs low | Prot-LC2 | -0.05 [-0.12; 0.02] | 0,19 |
| Temperature (previous month) - postnatal |  | Prot-LC2 | 0 [-0.04; 0.04] | 0,93 |
| Temperature (t1) - prenatal |  | Prot-LC2 | 0.01 [-0.03; 0.04] | 0,75 |
| Total fat intake - postnatal | Medium vs low | Prot-LC2 | -0.05 [-0.12; 0.02] | 0,15 |
| Total fat intake - postnatal | High vs low | Prot-LC2 | 0.01 [-0.06; 0.08] | 0,8 |
| Traffic density on major road (home) - postnatal | Yes | Prot-LC2 | -0.04 [-0.11; 0.03] | 0,31 |
| Traffic density on major road - prenatal | Yes | Prot-LC2 | -0.01 [-0.07; 0.05] | 0,69 |
| Traffic density on nearest road (home) - postnatal |  | Prot-LC2 | -0.01 [-0.06; 0.03] | 0,56 |
| Traffic density on nearest road - prenatal |  | Prot-LC2 | -0.01 [-0.04; 0.02] | 0,53 |
| UV - Vit.D (month) - postnatal |  | Prot-LC2 | -0.01 [-0.06; 0.03] | 0,56 |
| Vegetables intake - postnatal | Medium vs low | Prot-LC2 | -0.04 [-0.11; 0.04] | 0,36 |
| Vegetables intake - postnatal | High vs low | Prot-LC2 | -0.01 [-0.07; 0.06] | 0,8 |
| Walkability - prenatal |  | Prot-LC2 | 0 [-0.05; 0.05] | 0,93 |
| Walkability index - postnatal |  | Prot-LC2 | -0.01 [-0.05; 0.04] | 0,71 |
| Water Brominated THMs - prenatal |  | Prot-LC2 | 0 [-0.04; 0.04] | 0,9 |
| Water Chloroform - prenatal |  | Prot-LC2 | 0 [-0.05; 0.05] | 0,94 |
| Water THMs - prenatal |  | Prot-LC2 | 0.01 [-0.03; 0.06] | 0,62 |
| Yogurt intake - postnatal | Medium vs low | Prot-LC2 | 0.03 [-0.04; 0.1] | 0,41 |
| Yogurt intake - postnatal | High vs low | Prot-LC2 | 0 [-0.07; 0.06] | 0,91 |
| Accessibility (bus lines 300m) - postnatal |  | WBC-LC1 | 0.01 [-0.05; 0.06] | 0,83 |
| Accessibility (bus lines 300m) - prenatal |  | WBC-LC1 | 0 [-0.06; 0.06] | 0,97 |
| Accessibility (bus stops 300m) - postnatal |  | WBC-LC1 | 0.01 [-0.05; 0.06] | 0,83 |
| Accessibility (bus stops 300m) - prenatal |  | WBC-LC1 | 0.01 [-0.03; 0.04] | 0,74 |
| Bakery products intake - postnatal | High vs low | WBC-LC1 | 0.02 [-0.05; 0.09] | 0,53 |
| Bakery products intake - postnatal | Medium vs low | WBC-LC1 | 0.02 [-0.04; 0.08] | 0,56 |
| Blue spaces (300 m) - postnatal | Yes | WBC-LC1 | 0 [-0.1; 0.1] | 0,97 |
| Blue spaces (300 m) - prenatal | Yes | WBC-LC1 | 0.12 [0.01; 0.22] | 0,03 |
| Bread intake - postnatal | High vs low | WBC-LC1 | -0.01 [-0.08; 0.06] | 0,78 |
| Bread intake - postnatal | Medium vs low | WBC-LC1 | 0 [-0.07; 0.06] | 0,95 |
| Breakfast cereals intake - postnatal | High vs low | WBC-LC1 | 0.03 [-0.04; 0.1] | 0,38 |
| Breakfast cereals intake - postnatal | Medium vs low | WBC-LC1 | 0.01 [-0.05; 0.08] | 0,75 |
| Built density (300m) - postnatal |  | WBC-LC1 | 0 [-0.03; 0.03] | 0,98 |
| Built density (300m) - prenatal |  | WBC-LC1 | 0 [-0.04; 0.03] | 0,81 |
| Caffeinated drinks - postnatal | High vs low | WBC-LC1 | 0.06 [-0.01; 0.12] | 0,09 |
| Caffeinated drinks - postnatal | Medium vs low | WBC-LC1 | 0.07 [-0.01; 0.14] | 0,1 |
| Cat at home - postnatal | Yes | WBC-LC1 | 0.01 [-0.06; 0.08] | 0,78 |
| Cereals intake - postnatal | High vs low | WBC-LC1 | 0.05 [-0.02; 0.11] | 0,16 |
| Cereals intake - postnatal | Medium vs low | WBC-LC1 | 0.02 [-0.05; 0.08] | 0,61 |
| Cereals intake - prenatal | Medium vs low | WBC-LC1 | -0.03 [-0.1; 0.03] | 0,32 |
| Cereals intake - prenatal | High vs low | WBC-LC1 | -0.02 [-0.08; 0.05] | 0,64 |
| Connectivity (300m) - prenatal |  | WBC-LC1 | -0.02 [-0.06; 0.02] | 0,37 |
| Connectivity density (300 m) - postnatal |  | WBC-LC1 | -0.01 [-0.05; 0.03] | 0,55 |
| Contact with family and friends - postnatal |  | WBC-LC1 | 0.05 [-0.08; 0.17] | 0,45 |
| Contact with family and friends - postnatal |  | WBC-LC1 | -0.02 [-0.07; 0.04] | 0,61 |
| Dairy products intake - postnatal | Medium vs low | WBC-LC1 | 0.02 [-0.05; 0.08] | 0,6 |
| Dairy products intake - postnatal | High vs low | WBC-LC1 | 0.01 [-0.06; 0.07] | 0,8 |
| Dog at home - postnatal | Yes | WBC-LC1 | 0.03 [-0.04; 0.11] | 0,39 |
| Exposure to smoke - postnatal |  | WBC-LC1 | -0.01 [-0.07; 0.04] | 0,61 |
| Facility density (300m) - postnatal |  | WBC-LC1 | 0.01 [-0.04; 0.05] | 0,77 |
| Facility density (300m) - prenatal |  | WBC-LC1 | 0.01 [-0.03; 0.05] | 0,66 |
| Facility richness (300m) - postnatal |  | WBC-LC1 | 0.01 [-0.03; 0.05] | 0,64 |
| Facility richness (300m) - prenatal |  | WBC-LC1 | 0.01 [-0.04; 0.05] | 0,75 |
| Family affluence score - postnatal |  | WBC-LC1 | -0.03 [-0.12; 0.07] | 0,56 |
| Family affluence score - postnatal |  | WBC-LC1 | -0.01 [-0.1; 0.08] | 0,87 |
| Family size - postnatal |  | WBC-LC1 | 0.02 [-0.01; 0.05] | 0,22 |
| Fastfood intake - postnatal | High vs low | WBC-LC1 | -0.04 [-0.13; 0.04] | 0,34 |
| Fastfood intake - postnatal | Medium vs low | WBC-LC1 | -0.02 [-0.08; 0.04] | 0,45 |
| Fish and seafood intake - postnatal | Medium vs low | WBC-LC1 | 0.03 [-0.03; 0.1] | 0,31 |
| Fish and seafood intake - postnatal | High vs low | WBC-LC1 | 0.03 [-0.04; 0.1] | 0,37 |
| Fish and seafood intake - prenatal | Medium vs low | WBC-LC1 | -0.06 [-0.13; 0] | 0,06 |
| Fish and seafood intake - prenatal | High vs low | WBC-LC1 | -0.02 [-0.08; 0.05] | 0,57 |
| Fruits intake - postnatal | High vs low | WBC-LC1 | 0.04 [-0.03; 0.11] | 0,23 |
| Fruits intake - postnatal | Medium vs low | WBC-LC1 | 0.02 [-0.05; 0.08] | 0,63 |
| Green spaces (300 m) - postnatal | Yes | WBC-LC1 | 0.04 [-0.03; 0.1] | 0,29 |
| Green spaces (300 m) - prenatal | Yes | WBC-LC1 | 0.04 [-0.03; 0.1] | 0,26 |
| Humidity (month) - postnatal |  | WBC-LC1 | -0.02 [-0.06; 0.02] | 0,43 |
| Humidity (t1) - prenatal |  | WBC-LC1 | -0.02 [-0.06; 0.02] | 0,41 |
| Indoor BTEX - postnatal |  | WBC-LC1 | -0.03 [-0.06; 0.01] | 0,1 |
| Indoor NO2 - postnatal |  | WBC-LC1 | 0.01 [-0.04; 0.05] | 0,71 |
| Indoor PM2.5 - postnatal |  | WBC-LC1 | -0.03 [-0.05; 0] | 0,03 |
| Indoor PMabsorbance - postnatal |  | WBC-LC1 | -0.01 [-0.04; 0.02] | 0,45 |
| Indoor benzene - postnatal |  | WBC-LC1 | -0.02 [-0.06; 0.01] | 0,19 |
| Inverse distance to nearest road - postnatal |  | WBC-LC1 | -0.03 [-0.06; 0.01] | 0,16 |
| Inverse distance to nearest road - prenatal |  | WBC-LC1 | -0.02 [-0.05; 0.01] | 0,23 |
| KIDMED score - postnatal |  | WBC-LC1 | 0.02 [-0.01; 0.05] | 0,19 |
| Land use (300m) - postnatal |  | WBC-LC1 | 0 [-0.04; 0.04] | 0,97 |
| Land use (300m) - prenatal |  | WBC-LC1 | 0.03 [0; 0.07] | 0,07 |
| Maternal smoking (active and ETS) - prenatal |  | WBC-LC1 | -0.02 [-0.07; 0.04] | 0,5 |
| Meat intake - postnatal | Medium vs low | WBC-LC1 | -0.04 [-0.11; 0.02] | 0,2 |
| Meat intake - postnatal | High vs low | WBC-LC1 | -0.01 [-0.08; 0.05] | 0,65 |
| Meat intake - prenatal | Medium vs low | WBC-LC1 | -0.02 [-0.08; 0.05] | 0,62 |
| Meat intake - prenatal | High vs low | WBC-LC1 | 0.01 [-0.06; 0.08] | 0,77 |
| Moderate and vigorous PA - postnatal |  | WBC-LC1 | -0.01 [-0.04; 0.03] | 0,61 |
| NDVI (100 m) - postnatal |  | WBC-LC1 | 0 [-0.04; 0.04] | 1 |
| NDVI (100 m) - prenatal |  | WBC-LC1 | 0 [-0.04; 0.05] | 0,91 |
| Organic food intake - postnatal | High vs low | WBC-LC1 | 0.03 [-0.04; 0.1] | 0,39 |
| Organic food intake - postnatal | Medium vs low | WBC-LC1 | -0.02 [-0.08; 0.04] | 0,59 |
| Other pets at home - postnatal |  | WBC-LC1 | 0.03 [-0.02; 0.09] | 0,27 |
| Outdoor NO2 (preg) - prenatal |  | WBC-LC1 | -0.01 [-0.04; 0.02] | 0,54 |
| Outdoor NO2 (year) - postnatal |  | WBC-LC1 | 0 [-0.05; 0.04] | 0,9 |
| Outdoor PM10 (preg) - prenatal |  | WBC-LC1 | -0.01 [-0.04; 0.03] | 0,63 |
| Outdoor PM10 (year) - postnatal |  | WBC-LC1 | 0.01 [-0.03; 0.04] | 0,76 |
| Outdoor PM2.5 (preg) - prenatal |  | WBC-LC1 | -0.02 [-0.06; 0.02] | 0,35 |
| Outdoor PM2.5 (year) - postnatal |  | WBC-LC1 | -0.01 [-0.03; 0.02] | 0,6 |
| Outdoor PMabsorbance (preg) - prenatal |  | WBC-LC1 | -0.01 [-0.05; 0.03] | 0,73 |
| Outdoor PMabsorbance (year) - postnatal |  | WBC-LC1 | 0.01 [-0.02; 0.05] | 0,41 |
| Parental smoking - postnatal |  | WBC-LC1 | -0.04 [-0.12; 0.05] | 0,44 |
| Parental smoking - postnatal |  | WBC-LC1 | 0.01 [-0.06; 0.07] | 0,83 |
| Population density - postnatal |  | WBC-LC1 | 0.02 [-0.01; 0.05] | 0,25 |
| Population density - prenatal |  | WBC-LC1 | 0.01 [-0.02; 0.04] | 0,41 |
| Potatoes intake - postnatal | High vs low | WBC-LC1 | -0.03 [-0.1; 0.05] | 0,52 |
| Potatoes intake - postnatal | Medium vs low | WBC-LC1 | 0.01 [-0.05; 0.07] | 0,69 |
| Pressure (t1) - prenatal |  | WBC-LC1 | 0.01 [-0.03; 0.05] | 0,68 |
| Processed meat intake - postnatal | Medium vs low | WBC-LC1 | 0.02 [-0.04; 0.08] | 0,53 |
| Processed meat intake - postnatal | High vs low | WBC-LC1 | 0 [-0.07; 0.07] | 0,92 |
| Ready made food intake - postnatal | High vs low | WBC-LC1 | 0.04 [-0.02; 0.11] | 0,22 |
| Ready made food intake - postnatal | Medium vs low | WBC-LC1 | -0.01 [-0.07; 0.06] | 0,86 |
| Road traffic load (100 m) - postnatal |  | WBC-LC1 | -0.03 [-0.08; 0.02] | 0,25 |
| Road traffic load (100 m) - prenatal |  | WBC-LC1 | -0.02 [-0.07; 0.03] | 0,44 |
| Sedentary behaviour - postnatal |  | WBC-LC1 | 0.01 [-0.02; 0.04] | 0,45 |
| Sleep duration - postnatal |  | WBC-LC1 | -0.02 [-0.05; 0.02] | 0,37 |
| Social participation - postnatal |  | WBC-LC1 | 0.04 [-0.04; 0.12] | 0,35 |
| Social participation - postnatal |  | WBC-LC1 | 0 [-0.06; 0.07] | 0,87 |
| Soda intake - postnatal | High vs low | WBC-LC1 | 0.07 [0; 0.14] | 0,04 |
| Soda intake - postnatal | Medium vs low | WBC-LC1 | 0.02 [-0.04; 0.08] | 0,57 |
| Sweets intake - postnatal | Medium vs low | WBC-LC1 | 0.08 [0.02; 0.15] | 0,01 |
| Sweets intake - postnatal | High vs low | WBC-LC1 | 0.04 [-0.03; 0.11] | 0,22 |
| Temperature (previous month) - postnatal |  | WBC-LC1 | 0.04 [0; 0.08] | 0,03 |
| Temperature (t1) - prenatal |  | WBC-LC1 | 0.01 [-0.02; 0.04] | 0,48 |
| Total fat intake - postnatal | High vs low | WBC-LC1 | -0.04 [-0.1; 0.03] | 0,27 |
| Total fat intake - postnatal | Medium vs low | WBC-LC1 | 0 [-0.07; 0.06] | 0,92 |
| Traffic density on major road (home) - postnatal | Yes | WBC-LC1 | 0 [-0.07; 0.06] | 0,96 |
| Traffic density on major road - prenatal | Yes | WBC-LC1 | -0.01 [-0.07; 0.04] | 0,68 |
| Traffic density on nearest road (home) - postnatal |  | WBC-LC1 | 0 [-0.04; 0.04] | 0,99 |
| Traffic density on nearest road - prenatal |  | WBC-LC1 | 0.01 [-0.02; 0.04] | 0,69 |
| UV - Vit.D (month) - postnatal |  | WBC-LC1 | 0.03 [-0.01; 0.07] | 0,16 |
| Vegetables intake - postnatal | High vs low | WBC-LC1 | 0.01 [-0.06; 0.07] | 0,84 |
| Vegetables intake - postnatal | Medium vs low | WBC-LC1 | 0 [-0.08; 0.07] | 0,89 |
| Walkability - prenatal |  | WBC-LC1 | 0.03 [-0.02; 0.07] | 0,24 |
| Walkability index - postnatal |  | WBC-LC1 | 0.01 [-0.04; 0.05] | 0,81 |
| Water Brominated THMs - prenatal |  | WBC-LC1 | -0.01 [-0.04; 0.03] | 0,77 |
| Water Chloroform - prenatal |  | WBC-LC1 | -0.02 [-0.07; 0.03] | 0,38 |
| Water THMs - prenatal |  | WBC-LC1 | -0.03 [-0.07; 0.01] | 0,15 |
| Yogurt intake - postnatal | Medium vs low | WBC-LC1 | -0.03 [-0.1; 0.04] | 0,36 |
| Yogurt intake - postnatal | High vs low | WBC-LC1 | -0.02 [-0.08; 0.04] | 0,51 |

This table gives the estimations for the associations between environmental exposures (Exposure, modality) and signatures, with their 95% confidence interval (CI) and p-values (p).

#### eTable 7 – Supplementary: description of the identified immune signatures from DNAm

| Analysis | R2 | N CpGs |
| --- | --- | --- |
| Main analysis | 0.1 % | 2 |
| DNAm uncorrected for WBC effects retained | 2.7 % | 11655 |
| Males only | 0.7 % | 3 |
| Females only | 0.3 % | 2 |
| Without cohort BIB | 0.8 % | 4480 |
| Without cohort INMA | 0.5 % | 2 |
| Without non-European | 0.4 % | 4154 |
| Without cohort RHEA | 0.4 % | 2 |
| Without cohort EDEN | 0.3 % | 2 |
| Without cohort MOBA | 0.2 % | 2 |
| Without cohort KANC | 0.1 % | 2 |
| Extreme values winsorized | 0.2 % | 2 |

This table describes the percentage of health score variance (R2) explained by DNAm-LC1 in the sensitivity analyses, ordered by the R2. It also includes the number of CpGs contributings significantly to DNAm-LC1 (N CpGs).

#### eTable 8 – Sensitivity analysis to the removal of WBC effects to DNAm: correlations between signatures

|  | Prot-LC1 | Prot_LC2 | Prot-LC3 | WBC-LC1 | WBC-LC2 | WBC-LC3 | DNAm-LC1 |
| --- | --- | --- | --- | --- | --- | --- | --- |
| Prot-LC1 | 1.00 | 0.00 | 0.00 | 0.20 | 0.06 | 0.00 | 0.19 |
| Prot_LC2 | 0.00 | 1.00 | 0.00 | -0.05 | -0.02 | -0.01 | -0.04 |
| Prot-LC3 | 0.00 | 0.00 | 1.00 | 0.01 | -0.01 | 0.06 | 0.00 |
| WBC-LC1 | 0.20 | -0.05 | 0.01 | 1.00 | 0.00 | 0.00 | 0.87 |
| WBC-LC2 | 0.06 | -0.02 | -0.01 | 0.00 | 1.00 | 0.00 | -0.05 |
| WBC-LC3 | 0.00 | -0.01 | 0.06 | 0.00 | 0.00 | 1.00 | 0.16 |
| DNAm-LC1 | 0.19 | -0.04 | 0.00 | 0.87 | -0.05 | 0.16 | 1.00 |

This table gives the pearson correlation coefficients between signatures built in the analysis where blood cell type effects was not corrected in DNAm (supplementary analysis) in the whole database (n=845).

### Complementary Methods

#### eMethod 1 - Exposome measures in pregnancy and childhood

Two main windows of exposure were considered, 1) a prenatal window including the pregnancy period, 2) a cross-sectional window in childhood including the simultaneous measure of exposome data, internal biomarkers and health outcomes.

A total of 31 prenatal and 60 childhood exposures were investigated in the study, including the outdoor exposome (air pollution, built environment, noise, green and blue space, and meteorological data), indoor air pollutants, water disinfection by products, lifestyle (exposure to tobacco smoking, diet and physical activity) and socio-economic factors. They were measured in diverse ways as described below.

The outdoor exposome was assessed through GIS information and existing land use regression models adjusted for data from regulatory monitors and remote sensing data (Maitre et al., 2018). In this particular study we analysed whole pregnancy and childhood levels of air pollution at the home address (the year average before follow-up), while first pregnancy trimester and childhood monthly levels of meteorological variables. Built environment was calculated in a 300 m buffer.

Lifestyle factors were assessed through standardized questionnaires: including the KIDMED questionnaires to assess Mediterranean diet (Serra-Majem et al., 2004), physical activity of the child, sleeping patterns of child, socioeconomic status (family affluence scale (Boyce et al., 2006), and subjective wealth), social capital of the family (Kritsotakis et al., 2008), exposure to environmental tobacco smoke, water consumption habits, cooking and heating methods at the home, cleaning products, bedroom location, noise perception, child’s use of mobile phones and other devices, use of green spaces, commuting behaviour, holidays and sun exposure, and puberty development of the child.

Concentrations of drinking water disinfection by products (DBPs) during pregnancy were estimated from water company concentration and habits obtained from questionnaire data. Data was not sufficiently complete to estimate child exposure to DBPs.

Indoor air concentrations of nitrogen dioxide (NO2), particulate matter <2.5μm (PM2.5), particulate matter absorbance (PMabs), benzene, and toluene, ethylbenzene, xylene (BTEX) were estimated through a prediction model that combined measurements in the homes of a subgroup of children with questionnaire data from the subcohort. Measurements of indoor NO2, benzene and TEX were conducted in the homes of 157 participants as part of the child panel study, which was nested within the HELIX subcohort in all cohorts except MoBa. Participants in the child panel study were followed for one week in two periods (approximately 6 months apart), and the last day of the first week coincided with the subcohort examination, including the completion of the main HELIX questionnaire.

#### eMethod 2 - Child biomarkers

##### Proteomics

Plasma protein levels were assessed using the antibody-based multiplexed platform from Luminex, at the Genomics Core facility at the Center for Genomic Regulation (CRG) (Spain). Three kits targeting 43 unique candidate proteins were selected (Thermo Fisher Scientifics, USA): Cytokines 30-plex (Catalog Number (CN): LHC6003M), Apoliprotein 5-plex (CN: LHP0001M) and Adipokine 15-plex (CN: LHC0017M). All samples were randomized and blocked by cohort prior measurement. For quantification, an 8- point calibration curve per plate was performed with protein standards provided in the Luminex kit and following procedures described by the vendor. Commercial heat inactivated, sterile-filtered plasma from human male AB plasma (Sigma-Aldrich, USA) was used as constant samples to control for intra- and inter-plate variability. Four control samples were added per plate. All samples, including controls, were diluted ½ for the 30-plex kit, ¼ for the 15-plex kit and 1/2500 for the 5-plex kit. Raw intensities obtained with the xMAP and Luminex system for each plasma sample were converted to pg/ml using the calculated standard curves of each plate and accounting for the dilutions made prior measurement. The percentages of coefficients of variation (CV%) for each protein by plate ranged from 3% to 36%. The limit of detection (LOD) and the lower and upper limit of quantification (LOQ1 and LOQ2, respectively) were estimated by plate, and then averaged. Only proteins with >30% of measurements in the linear range of quantification were kept in the database and the others were removed. Seven proteins were measured twice (in two different multiplex kits). We kept the measure with higher quality. The 36 proteins that passed the quality control criteria mentioned above were log2 transformed (Vives-Usano et al., 2020). Then, the plate batch effect was corrected by subtracting the plate specific average for each protein minus the overall average of all plates for that protein. After that, values below the LOQ1 and above the LOQ2 were imputed using a truncated normal distribution implemented in the truncdist R v1.0-2 package (Nadarajah & Kotz, 2006). Twenty samples were excluded due to having ten or more proteins out of the linear range of quantification.

##### Blood DNA methylation

DNA was obtained from children’s peripheral blood (buffy coat) collected in EDTA tubes. DNA was extracted using the Chemagen kit (Perkin Elmer, USA) in batches of 12 samples within each cohort. DNA concentration was determined in a Nanodrop 1000 UV-Vis Spectrophotometer (Thermo Fisher Scientific, USA) and also with Quant-iTTM PicoGreen dsDNA Assay Kit (Life Technologies, USA). DNA extraction was repeated in around 8% of the blood samples as the DNA quantity or quality of the first extraction was low. Less than 1.5% of the samples were finally excluded due to low quality.

DNA methylation was assessed with the Infinium HumanMethylatio450 beadchip (Illumina, USA) at the University of Santiago de Compostela – Spanish National Genotyping Center (CeGen- USC) (Spain). 700 ng of DNA were bisulfite-converted using the EZ 96-DNA kit (Zymo Research, USA) following the manufacturer’s standard protocol. All samples of the study were randomized considering sex and cohort. In addition, each plate contained a HapMap control sample and 24 HELIX inter-plate duplicates were included. After an initial inspection of the quality of the methylation data with the MethylAid v1.8.0 R package15, probes with a call rate <95% based on a detection p-value of 1E-16 and samples with a call rate <98% were removed. Samples with discordant sex were eliminated from the study as well as duplicates with inconsistent genotypes and samples with inconsistent genotypes respect to existing genome-wide genotyping array data. Methylation data was normalized using the functional normalization method with prior background correction with Noob17. Then, some probes were filtered out: control probes, probes to detect single nucleotide polymorphisms (SNPs), probes to detect methylation in non-CpG sites, probes located in sexual chromosomes, cross hybridizing probes (Chen et al., 2013), probes containing a SNP at any position of the sequence with a minor allele frequency (MAF) >5% and probes with a SNP at the CpG site or at the single base extension (SBE) at any MAF in the combined population from 1000 Genomes Project. CpGs were annotated with the IlluminaHumanMethylation450kanno.ilmn12.hg19 v0.6.0 R package20.

Additional pre-treatment of CpGs was done. 1) The most extreme values (>95th percentiles, <5th percentiles) were processed using winsozisation for each CpGs. It consists in changing the 5% highest values to the 95th percentile, and the 5% lowest values to the 5th percentile, for a given site. 2) We corrected remaining technical batch effects and blood cell composition by using a residualization analysis. Briefly, we fitted a linear regression model between each CpGs and technical predictors (batch, array) plus white blood cell type percentage (which is one of the biggest determinants of methylation levels). For this step, neutrophiles were removed from the predictors to avoid multicolinearity. Then, we extracted the residuals of the linear regression and used it as corrected methylation values. We chose to remove the white blood composition effect from DNA methylation in the main analysis, and study it in more details as a separate layer. We also performed a secondary analysis in which white blood composition effect was not removed in the methylation data, i.e. we residualized on technical factors only. 3) We excluded probes for CpGs that did not reach a 62.5% interclass correlation coefficient (ICC) to minimize technical variability, based on a previous analysis with technical replicates (Bose et al., 2014). 4) We filtered out the CpGs with no association with the exposome based on the study of Maitre (Maitre et al., 2022).

As a result, a total of 20,386 CpG sites were selected from the 480,071 sites initially measured.

##### White Blood Cell composition

A total of twelve white blood cell proportions (neutrophils, eosinophils, basophils, monocytes, naïve and memory B cells, naïve and memory CD4 + and CD8 + T cells, natural killer, and T regulatory cells) were estimated using the Enhanced Salas algorithm from raw methylation data of WBC (see method section on DNA methylation above).

We excluded 44 samples due to poor DNA methylation quality control (see more details in the DNA methylation section above). Then, we pre processed outliers and zero. We identified absolute zeros, very small values close to zero (we used a threshold of <1e-4), or negative values that could act as influential points in the analyses. Then, these values were imputed with the robComposition R package (Templ et al., 2011; Martín-Fernández et al., 2012). This method uses robust rather than classical regressions in the iterative scheme. Robust regression automatically down-weights outlying observations, i.e., observations that are outliers in either the space of the predictors or the residual space.

#### eMethod 3 - Multi-Domain Health Score

For each health domain (cardiometabolic, respiratory/allergy and mental), a sub-score was created. Following the approach of Eisenmann (Eisenmann, 2008), subscores were defined as the sum of z-scores or multiple factor analysis (applying equal weighting to predefined groups of variables). All of the three sub-scores were built such that a high score means the child is in good health on this specific domain.

The three sub-scores were scaled and aggregated into one multi domain health score by taking their mean. By construct, the health score is low for children with conjointly low-to-moderate cardiometabolic, respiratory/allergy and mental health in children, as well as for children highly affected in one health domain while no or moderately affected for the other two.

##### Cardiometabolic subscore

**Cardiometabolic outcomes** (Vrijheid et al., 2020; Warembourg et al., 2019)

Blood pressure was measured during the clinical examination using a standardized protocol: after 5 minutes of rest in sitting position, 3 consecutive measurements were taken by oscillometric device (OMRON 705-CPII) with one-minute time intervals between them, in a predefined posture and in preference in the right arm. Adequate cuff sizes were chosen with respect to each child’s arm length and circumference. Systolic (SBP) and diastolic (DBP) blood pressures from each measurement have been recorded and the mean of the second and the third measurements was calculated and used in this analysis. During the subcohort examination, the waist circumference was measured as an indicator of visceral fat in duplicate to the nearest 0:1 cm in a standing position, at the high point of the iliac crest at the end of a gentle expiration, with the use of a measuring tape (Seca 201; Seca Corporation). Also, height and weight were measured using regularly calibrated instruments. For all measures, we used common standardized protocols and the same instruments across the cohorts. Concentrations (mg/dL) of triglycerides and high-density lipoprotein (HDL) cholesterol was measured in child nonfasting serum using homogeneous enzymatic colorimetric methods on a Modular Analytics System from Roche Diagnostics GmbH Mannheim and according to the manufacturer’s instructions 4. Insulin levels were measured in serum using human adipokine 15-plex magnetic panels 5.

**Cardiometabolic subscore**

Continuous health parameters were transformed in z-scores, using Generalize Additive Model for Location, Scale and Shape (GAMLSS) (Rigby et al., 2019) to standardize on covariates and approach normality. For blood pressure, a standardization for height was added (Barba et al., 2014).

As used previously in the Helix population, the cardiometabolic sub-score was defined as (-z waist circumference) + (- z insulin) + (z HDL cholesterol – z triglycerides)/2 + (-z systolic BP – z diastolic BP)/2 (Ahrens et al., 2014; Stratakis et al., 2020).

##### Respiratory and allergic subscore

**Respiratory and allergic outcomes** (Agier et al., 2019; Granum et al., 2020)

Lung function was measured by a spirometry test (EasyOne spirometer; NDD [New Diagnostic Design], Zurich, Switzerland), by trained research technicians using a standardised protocol. The child, sitting straight and equipped with a nose clip, was asked to perform at least six manoeuvres (if possible). Data from unacceptable manoeuvres as a result of errors, including hesitation or false starts, cough, variable efforts, glottis closure, early termination, and leaks, were not retained by the technicians. The protocol required that at least three acceptable manoeuvres were obtained, and that they were reproducible, defined as a difference of less than 200 mL between the two highest values for forced vital capacity (FVC) and Forced Expiratory Volume in one second (FEV₁) taken from the acceptable manoeuvres. We then applied the following validation criteria on the spirometer curves retained by the technicians. We defined a manoeuvre as acceptable if there was no hesitation or false start (defined as a ratio of backward extrapolated volume [BEV] to FVC of <5% or a BEV of <100 mL if FVC was <1000 mL) and if the forced expiratory time was in an acceptable range (>1·5 s and <10 s). The two highest values for FEV₁ taken from acceptable forced expiratory manoeuvres could not vary by more than 150 mL or by more than 5% from the second FEV₁. To address the efficiency of the FEV₁ data cleaning, the 243 examinations from the INMA cohort (185 of which were included in the HELIX study) were further investigated by trained investigators who looked at the shape of the curves; for 192 (79%) of the 243 examinations, the same curve was selected, and for the remaining 51 (21%), the Pearson correlation between the FEV₁ of the two different curves was 0·96. We used the reference equations estimated by the Global Lung Initiative (Stanojevic et al., 2017) for computing the FEV₁ percent predicted values (ie, values standardised by age, height, sex, and ethnicity of the patient) and FEV₁ Z scores. After excluding extreme values (ie, FEV₁ <60% or >140%, which were probably due to measurement error in our young population), we selected the greatest FEV₁ value at an individual level among all accepted curves, hereafter referred to as FEV₁%. For this analysis, we used FEV₁%. as our main measure of the lung function. Information on the asthma and allergy-related outcomes were obtained through questions adapted from the International Study on Asthma and Allergy in Childhood (ISAAC) (Asher et al., 1995) during the interview with the mother at the follow-up examination. “Has your child ever been diagnosed by a doctor as having asthma?” (hereafter called asthma); “Has your child ever been diagnosed by a doctor as having eczema or atopic dermatitis or neurodermatitis?” (hereafter called eczema); “Has your child ever had an allergic reaction to food, diagnosed by a doctor?” (hereafter called food allergy); “In the last year, has your child had problems with sneezing, or a runny, or blocked nose when he/she did not have a cold or the flu?” (hereafter called rhinitis).

**Respiratory/allergic subscore**

The respiratory/allergy sub-score was defined as the first principal component from a multiple factorial analysis, chosen because it accommodates mixed-type variables. This analysis grouped asthma, allergies, eczema, and rhinitis in one category and FEV1% predicted in another. The coordinates and contributions of each variable to the first component are available in Amine et al., 2023 (supplementary materials, eTable 7). Asthma and allergy-related variables had negative coordinates, while lung function (FEV1) also had a positive coordinate, indicating that better lung function and fewer allergy-related conditions correspond to a higher respiratory/allergy sub-score.

##### Mental/Neurodevelopmental subscore

**Neurodevelopmental outcomes**

Trained fieldwork technicians measured the fluid intelligence using a computer-based test which is the Raven Colour Progressive Matrix™ 9 (CPM). The CPM comprised a total of 36 items and we used the total number of correct responses as the outcome. A higher CPM scoring indicates better fluid intelligence. Fluid intelligence is the ability to solve novel reasoning problems and depends only minimally on prior learning. All examiners were previously trained following a standardized assessment protocol by the study expert psychologist. Furthermore, during the pilot phase, a coordinator visited each cohort site and checked for any potential error committed by the previously trained examiners.

Parents completed questionnaires related to child’s behaviour, including the child behaviour checklist (CBCL) and the Conner rating scale’s, within a week before the follow-up visit at 6–12 years of age. The 99-item CBCL/6–18 version for school children was used to obtain standardized parent reports of children’s problem behaviours, translated and validated in each native language of the participating six cohort populations (Neo et al., 2021). The parents responded along a 3-point scale with the code of 0 if the item is not true of the child, 1 for sometimes true, and 2 for often true. The internalizing score includes the subscales of emotionally reactive and anxious/depressed symptoms, as well as somatic complaints and symptoms of being withdrawn. The externalizing score includes attention problems and aggressive behaviours. In addition, an ADHD index based on the short form of the Conners’ rating scales of 27 items provided information on inattention and hyperactivity symptoms (Gurley, 2011). The internal consistency (Cronbach’s alpha) of each of the study scales was >0.80. All outcomes were analyzed as raw count scores.

**Neurodevelopmental subscore**

Continuous health parameters were transformed in z-scores, using Generalize Additive Model for Location, Scale and Shape (GAMLSS) (Rigby et al., 2019) to standardize on covariates and approach normality.

The mental sub-score was derived as the first principal component from a multiple factorial analysis. This method was chosen because, to our knowledge, no mental score based on z-scores has been reported in the literature. The factorial analysis grouped the z-scores of behavioral parameters (ADHD, internalizing, externalizing) in one category and the cognitive parameter (fluid intelligence) in another. The coordinates and contributions of each variable to the first component can be found in Amine, 2023 (supplementary materials). Behavioral scores had negative coordinates, while the cognitive score had a positive coordinate, indicating that better cognition and fewer behavioral difficulties correspond to a higher neurodevelopmental sub-score.

#### eMethod 4 - Pre-processing of the exposures and covariates

##### Transformation of exposures/covariates

For each variable, the optimal transformation to approach normality was chosen using a Box-Cox power transformation approach and described in eTable 1. When normality could not be achieved by transformation, the variable was categorized. To improve comparison between exposures, estimates reported for continuous variables are expressed as an increase in interquartile range (IQR) (see eTable 1).

##### Imputation process

To prevent losing information and introducing potential selection biases, missing values of exposures and covariates were imputed. When possible, we relied on the multiple imputation procedure, which is a commonly used and accepted method to deal with missing data (Stuart et al., 2009). Multiple imputation provides valid inferences under the missing at random (MAR) assumption, which assumes that missing data are associated to observed variables and not to unobserved information. In multiple imputation, missing values are imputed stochastically several times. Imputing missing values several times allows the quantification of the uncertainty in results associated with imputation, and to account for this uncertainty in the final standard errors, confidence intervals and p-values.

For the imputation process, continuous variables should have a normal distribution. As mentioned above, skewed exposure variables were transformed to achieve normality or categorized if no transformation worked. The distributions of all transformed variables were examined to make sure that transformations did not lead to extreme/influential observations. In cases of variables with zeros that required a log transformation, a constant value was added to the variable as the log of zero is minus infinity. The constant value was chosen to minimize the skewness of the resulting variable. Exposures with more than 30% of missing values were excluded from the process.

Missing values of exposures and adjustment variables were imputed using the method of chained equations (White et al., 2011), using the mice package in R (Buuren & Groothuis-Oudshoorn, 2011). The large number of variables involved in the HELIX analyses implies that not all exposures can be used as predictors in imputation models, as imputation models would be too large, and therefore reduced imputation models had to be specified. It is recommended that imputation models include between 10 and 25 variables (Buuren & Groothuis-Oudshoorn, 2011). We used the quickpred function to reduce the number of predictors. With this function, potential predictors for a given variable var1 were restricted to those that had: 1) an absolute correlation greater than 0.2 with var1 or with the binary variable indicating if var1 is missing (mincor=0.2); and 2) a proportion of non-missing observations greater than 40% among the observations with missing values in var1 (minpuc=0.4). Values for mincor and minpucwere tuned to have a number of predictors between 5 and 25 for all variables. If some variables continued to have too many predictors, they were reduced manually. In addition, we forced into the imputation models cohort and one representative outcome variable of each the main groups of health outcomes that will be subsequently analysed in the HELIX project (namely asthma as representative of respiratory health; z-score of BMI as representative of cardio metabolic outcomes; and number of correct responses in RAVEN test and total problems in CBCL as representative of neurodevelopment). Those variables were included with the include option. With the procedures described, the imputation process still produced some errors, which were solved on a case-by-case basis. Mostly, this consisted in removing from the predictor set of a given variable highly correlated variables. The method of predictive mean matching was used for all continuous exposures.

Twenty imputed datasets were created, although only the first imputed dataset was used in this paper. After imputation, the following diagnostics were conducted.

All variables with missing values were inspected. In particular, the imputed and non-missing observations were compared using density plots and stripplots of Van Buuren (Buuren & Groothuis-Oudshoorn, 2011). These types of comparison were only done when there were more than 5% of the observations with missing values. Numerically, variables were flagged if they have 1) an absolute difference between means of the observed and imputed values greater than 2 standard deviations; or 2) a ratio of variances of the observed and imputed values that is less than 0.5 or greater than 2 (Stuart et al., 2009). For categorical variables, those with a significant chi-squared test between imputed and non-imputed were examined. If variables were flagged and imputations seem implausible, the predictors included in the imputation model for that variable were changed to those plausible imputations were generated.

Under the missing at random hypothesis, imputing the outcomes would not benefit the model compared to an analysis on complete cases (Little, 1992). Therefore, all participants with at least one missing value on the health score and the immune biomarkers were removed, leading to the inclusion of 845 mother-child pairs.

#### eMethod 5 - Exposome-Biomarkers-Health association study

##### Step 1: Multiblock model – Identification of immune signatures predicting the health score

A multiblock model was employed to examine the combined impact of multi-omic immune biomarkers on the health score, recognizing that they influence health collectively rather than independently. This approach aimed to identify inflammation profiles associated with the health score.

**RGCCA**

Regularized Generalized Canonical Correlation Analysis (RGCCA, R package) was used to estimate latent components (signatures) for each data block (biological layer) that are strongly correlated with the health score and potentially with signatures from other blocks, as connections are defined across all blocks (Tenenhaus & Tenenhaus, 2011). While a similar model could be built using the mixOmics package (“multiblock PLS”), RGCCA was preferred because mixOmics does not allow parameter tuning via cross-validation or loading estimation via bootstrap. To account for confounders, biomarkers and the outcome were adjusted for covariates beforehand using linear regression (residualization approach).

**Model implementation**

The three blocks used were: 1) white blood cell composition, 2) plasma proteins, 3) DNA methylation.

1) Cross-validation was used to tune the number of components (ranging from 1 to 3 per block) and the sparsity parameter (applied only to DNA methylation). Default RGCCA settings were used for other parameters. Optimal hyperparameters were selected based on the model's predictive performance (RMSE), assessed through a linear regression predicting the health score from the estimated signatures. The final model was then built using the optimized parameters on the full dataset. To ensure consistency, signatures were multiplied by -1 if they were negatively associated with the health score, so that all signatures were associated with better health outcomes.

2) Prediction accuracy was evaluated through cross-validation. For each Kth fold, R² and p-values were reported from linear regression models to assess the ability of immune signatures (individually and collectively) to predict the health score.

3) Bootstrap resampling was used to estimate confidence intervals and p-values for the loadings, representing biomarker contributions to the signatures.

4) In the full dataset, associations between the signatures and the health scores (multi-domain health score and subscores) were estimated using univariate linear regression models. Covariate adjustment was unnecessary, as the variables had already been pre-corrected before running RGCCA.

##### Step 2: Association between environmental exposures and identified immune signatures

Associations between environmental exposures and the immune signatures were assessed using univariate linear regression models. Covariate adjustment was not necessary, as exposures, biomarkers, and outcomes had already been pre-corrected through residualization before running RGCCA.

##### Supplementary analyses

Stratified analyses were conducted by child sex to explore possible sex-specific effects.

In addition, to ensure the robustness of the findings, several analyses were conducted:

1) A leave-one-cohort-out approach was applied to ensure that results were not driven by a single cohort.

2) The 4% most extreme outcome values (2% lowest and 2% highest) were removed to mitigate the influence of potential outliers.

3) An analysis excluding non-European children was performed to account for potential ethnic-specific effects.

4) An analysis using the DNA methylation for which the WBC composition effect was not removed during the pre-processing step.

We also performed our results to a more traditional “Meet in the middle” approach using univariate regression models for the exposome-biomarker and biomarker-health score sides (Chadeau-Hyam et al., 2011). For this step, pre-filtering of CpGs based on exposome association, as done in the multiblock model, was not applied. For the exposome-biomarker analysis, results from the Helix population were extracted using the ExWAS catalogue (https://helixomics.isglobal.org/), with new results obtained for WBC composition, which was not included in the catalogue. For the biomarker-health score analysis, univariate regressions were conducted for each biomarker with the health score, adjusting for the same covariates used in other analyses. Significant associations were defined as having a p-value below 1% for DNA methylation and 5% for proteins and cell types. Corrected p-values for multiple testing were reported. Within each omics layer, p-values were adjusted for the number of molecular features specific to each platform. For methylation, we applied the False Discovery Rate (FDR) using the Benjamini-Hochberg (BH) method (Benjamini & Hochberg, 1995). For other omics, the effective number of tests (ENT) was calculated, and nominal p-values (0.05) were divided by this number (Li et al., 2012).
